# Spatiotemporal Patterns and Climate-Driven Forecasting of Scrub Typhus: Evidence from South India

**DOI:** 10.64898/2026.03.18.26348670

**Authors:** R Bithia, Muzaffar Ahmad Dar, Solomon D Cruz, CL Biji, Manglam Goutam Sinha, Aman Picardo, Aadithya Hrudhay Anand, Bikash Keshari, P Prithviraj, Surendar Manickam, C George Priya Doss, Karthik Gunasekaran, John Antony Jude Prakash

## Abstract

Scrub typhus remains a persistent public health concern with strong spatial and temporal variability. This study analyses the spatio-temporal distribution, clustering patterns, and forecasting of scrub typhus across five districts, Chittoor, Ranipet, Tirupattur, Vellore, and Tiruvannamalai, using long-term surveillance data from May 2005 to May 2024. We applied spatio-temporal exploratory analysis to identify trends, seasonal behaviour, and inter-district heterogeneity in disease incidence. Hotspot analysis was conducted using the Getis-Ord Gi* statistics to detect statistically significant hotspots and coldspot clusters and examine their evolution over time. To support decision-making, we developed statistical, machine learning (ML), and deep learning (DL) based forecasting models using monthly scrub typhus and climatic features. Root mean square error (RMSE), and R-square error (*R*^2^) evaluation metrics are used to compare the performance of the prediction model. Scrub typhus shows clear and recurring seasonal peaks across all five districts, and incidence increases are associated with precipitation, dew point, relative humidity, and vegetation cover. Temperature shows a strong negative correlation, while relative humidity and normalized difference vegetation index (NDVI) show strong positive correlations in all districts. Hotspot analysis identifies Vellore and Chittoor as persistent core transmission zones, with weaker clustering in surrounding districts. Forecasting results indicate that model performance varies by location. The results reveal persistent hotspots, clear seasonal signals, and short-term forecasts across districts. This integrated spatiotemporal and forecasting framework provides actionable insights for targeted surveillance and timely intervention strategies to control scrub typhus.

## 1. Introduction

Scrub typhus is an acute febrile illness caused by *Orientia tsutsugamushi* and transmitted to humans through the bite of infected chiggers, larvae of trombiculid mites [1]. The disease commonly presents with fever, headache, myalgia, and, in some cases, a characteristic eschar at the mite (chigger) bite site [2]. However, the absence of eschar in many patients often leads to diagnostic uncertainty, as scrub typhus can closely mimic other acute febrile illnesses, including dengue, malaria, leptospirosis, and enteric fever [3]. Laboratory confirmation, most often through IgM ELISA testing, is therefore essential before initiating appropriate treatment in clinically suspected cases [4]. In India, scrub typhus has emerged as a primary public health concern, accounting for a significant proportion of undifferentiated febrile illnesses. Around one-fifth of affected patients may progress to severe disease, requiring intensive care due to complications such as multiorgan dysfunction [5].

The transmission dynamics of scrub typhus are closely linked to environmental and climatic conditions that influence the survival and distribution of vector mites [6,7]. Changes in temperature, rainfall, humidity, vegetation cover, and land-use patterns can alter human-vector contact, thereby shaping disease incidence [6, 8, 9]. Vector-borne infections such as scrub typhus are susceptible to climate variability, ecological disruption, and expanding human activity, making them increasingly unpredictable in both space and time [6–9]. Understanding how these factors interact at a regional level is essential for effective disease surveillance, risk assessment, and targeted public health interventions.

In recent years, a growing body of research has examined the ecological and climatic drivers of scrub typhus using advanced statistical and spatial methods. Various analytical approaches, including univariate and multivariate logistic regression, have been employed to identify predictors of disease severity and clinical outcomes [10]. Studies from Nepal and other endemic regions have demonstrated the utility of integrating meteorological variables, vegetation indices, topography, and land-cover data to forecast scrub typhus risk, with ML models such as random forest and maximum entropy showing promising performance [11]. Geographic information systems (GIS) have further contributed to mapping disease distribution and identifying high-risk areas based on environmental suitability [12]. Long-term analyses using historical case data and vector distribution maps have also provided valuable insights into the evolving geographic range of scrub typhus [13, 14]. More recently, ML techniques, including artificial neural networks (ANN), have been applied to capture non-linear relationships between meteorological variables and scrub typhus incidence. These studies have highlighted the influence of minimum temperature, relative humidity, rainfall, and wind speed on disease transmission and seasonal trends [15, 16]. Environmental changes, such as deforestation, have also been strongly associated with increased scrub typhus risk, as demonstrated by satellite-based analyses and Bayesian modelling [17]. Hybrid spatio-temporal models developed in regions such as Korea and China further underscore the value of combining climatic, geographic, and demographic data to improve outbreak prediction and early warning systems [18].

In India, research on scrub typhus has increasingly emphasized regional patterns and seasonal variability, particularly in southern states where the disease burden remains high. In Tamil Nadu, hospital-based and regional studies have consistently identified scrub typhus as a leading cause of acute undifferentiated febrile illness, with cases reported both from urban and rural settings [19,20]. Several studies from South India have documented a distinct seasonal trend, with increased incidence during cooler and post-monsoon periods, highlighting the influence of local climatic conditions on transmission dynamics [21]. A notable longitudinal study from a tertiary care center in Vellore analysed 15 years of scrub typhus surveillance data and identified significant associations between meteorological variables such as temperature, rainfall, and humidity, and disease incidence using negative binomial regression models [22]. While this work provided important insights into climate-disease relationships, its scope was limited to a single district and a specific modelling approach, underscoring the need for validation across an extended time period, broader geographic settings, and alternative predictive frameworks.

Building on this earlier work, the present study uses the same core dataset, augmented by an additional 5 years of data, enabling a more robust evaluation of long-term trends in scrub typhus incidence. Furthermore, the analysis is expanded beyond Vellore to include four neighbouring districts, Ranipet, Chittoor, Tiruvannamalai, Tirupattur, and Vellore, thereby capturing spatial variability within an endemic region of Tamil Nadu. In addition to commonly examined meteorological variables, we include a broader set of climate and environmental features, such as dew point, wind speed, skin temperature, and NDVI, to better represent conditions that influence vector ecology and disease transmission. To further enhance predictive performance, the analysis incorporates time-series decomposition components and engineered features that capture seasonal patterns, trends, and lagged effects. Using this enriched feature set, we systematically evaluate classical time-series models (ARIMA, SARIMA), machine-learning approaches, and deep-learning models for predicting scrub typhus incidence. By integrating extended temporal coverage, multidistrict spatial analysis, and a richer representation of climatic and environmental drivers, this study aims to improve district-level risk prediction and provide practical tools to support early warning systems and targeted public health interventions in scrub typhus-endemic regions. Figure 1 shows the detailed workflow of the proposed study for the construction of the scrub typhus modelling.

**Figure 1:**
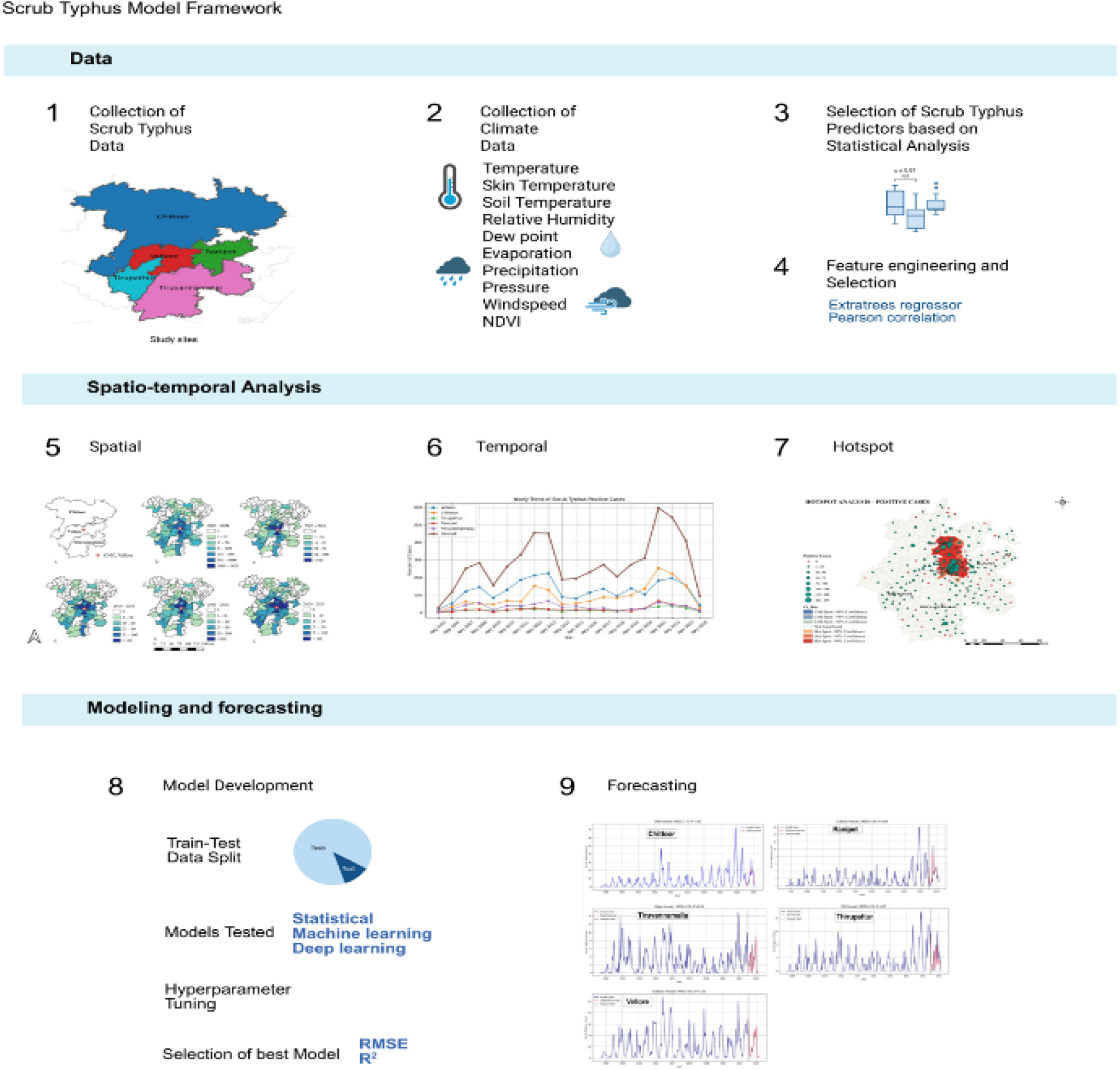
The Scrub Typhus modelling framework illustrates spatial analysis, temporal modelling, and predictive projections of Scrub Typhus incidence.

## 2. Methods

### 2.1 Overview of the study area

The Vellore–Tiruvannamalai region, including Tirupattur, Ranipet, and adjoining Chittoor, forms a contiguous belt at the foothills of the Eastern Ghats as shown in Figure 2. These regions were selected as the study area considering their persistently elevated scrub typhus incidence rates over the 15-year period (May 2005 to May 2024). The region lies in a tropical wet–dry zone, and its climate is shaped by the monsoon. Rain comes in two main seasons: the southwest monsoon from June to September and the northeast monsoon from October to December. Together, these bring an annual rainfall of about 900 to 1100 millimetres. The approximate geographic ranges of these districts are as follows: Chittoor: 12°45′00″ to 13°45′00″ North latitude and 78°50′00″ to 79°40′00″ East longitude, Ranipet: 12°93’20.63’’ to 12°55’55.42’’ North Latitude and 79°33’66’’ to 79°20’47’’ East Longitude, Tirupattur: 12°29’21.77’’ to 12°49’83.51’’ North Latitude and 78°34’45’’ to 78°56’41’’ East Longitude, Tiruvannamalai: 12° 00’00’’to 12° 52’30’ North latitude and East Longitude 78° 39’30’’ to 79° 45’36, Vellore: 78°50′–79°30′ E, 12°35′–13°10′N 12°15’23’’to 13°12’32’’ N Latitude, 78°24’16’’ to 79°54’56’’E Longitude.

**Figure 2:**
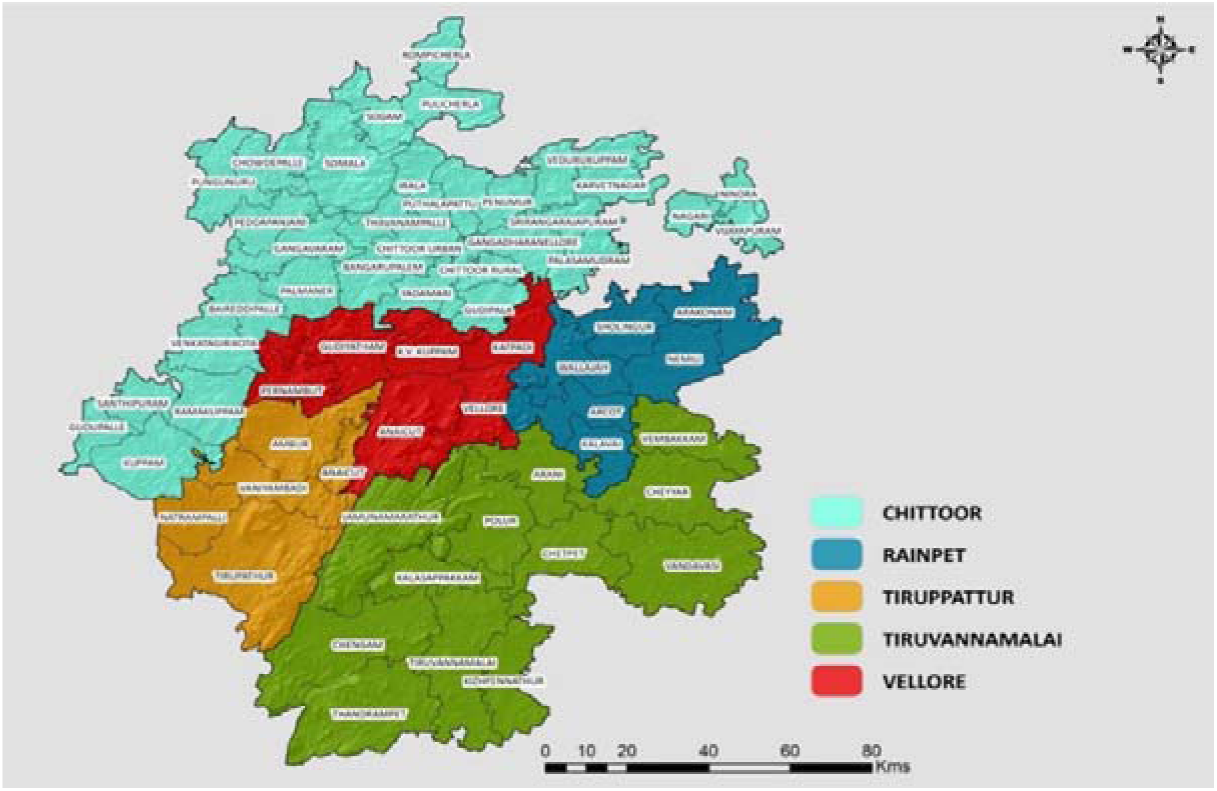
Geographical location of five selected districts in Tamil Nadu, India for scrub typhus modelling

### 2.2 Data sources

#### 2.2.1 Scrub typhus surveillance data

Scrub typhus-positive data for 15 years (May 2005 – April 2024) were retrieved from electronic medical records. A scrub typhus case was defined as an individual with acute undifferentiated febrile illness (AUFI) who tested positive for scrub typhus by IgM ELISA, tested negative for malaria and dengue, and had a negative blood culture. The scrub typhus serology was performed with appropriate IQA in the Immunology Laboratory, Department of Clinical Microbiology (ISO 15189 accredited lab), CMC, Vellore using an automated ELISA workstation (Euroimmun Analyzer I, Euroimmun AG, Lübeck, Germany).

#### 2.2.2 Climatic data

The study used two primary datasets: ERA5 (https://developers.google.com/earthengine/datasets/catalog/ECMWF_ERA5_LAND_DAILY_AGGR) and MOD13A2 (https://developers.google.com/earth-engine/datasets/catalog/MODIS_061_MOD13A2) vegetation indices. ERA5 Land provides a highresolution climate reanalysis of land surface and meteorological variables by integrating model simulations with global observations under physical consistency constraints. It offers long-term, temporally continuous climate information with up to 50 variables available through the Climate Data Store at an approximate spatial resolution of 11 km. Vegetation dynamics were derived from the MOD13A2, which provides the NDVI. It ensures continuity with NOAA AVHRR-based vegetation records and serves as an indicator of vegetation density and surface moisture. The dataset is available at a spatial resolution of 1000 m. Both datasets were processed and analysed using the Google Earth Engine (GEE) cloud platform, enabling large-scale geospatial computation and integration.

Based on disease burden and climatic variability, five districts—Chittoor, Ranipet, Tirupattur, Vellore, and Tiruvannamalai—were selected for analysis. Climatic variables hypothesized to influence scrub typhus transmission were collected for the corresponding study period. The extracted variables included relative humidity (%), dew point (°C), evaporation (mm), precipitation (mm), atmospheric pressure (hPa), skin temperature (°C), soil temperature (°C), maximum temperature (°C), mean temperature (°C), minimum temperature (°C), and normalized difference vegetation index (NDVI, % area). Climatic data were obtained at daily resolution and subsequently aggregated to match the temporal frequency of the scrub typhus surveillance dataset.

The scrub typhus surveillance dataset and the corresponding climatic dataset were available at a daily temporal resolution. To ensure temporal consistency and to enable monthly forecasting, both health and climate observations were aggregated to a monthly scale at the district level. Let *d* denote a district and *t* denote a calendar month. For meteorological variables representing intensity (dew point temperature, atmospheric pressure, NDVI area), relative humidity, skin temperature, soil temperature, and near-surface air temperature indices), monthly averages were computed as

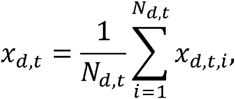

where *x_d,t,i_* is the daily value on day *i* in month *t* for district *d*, and *N_d,t_* is the number of available

daily observations in that month. In contrast, accumulation-type variables such as evaporation and precipitation were aggregated using monthly totals:

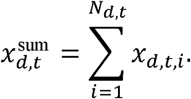

Scrub typhus and climatic datasets were checked for missing values, inconsistencies, and outliers. Missing climate values, when present, were imputed using propagation imputation, while missing scrub typhus values were treated as zero based on reporting context. All numeric features were scaled using standardization prior to applying modelling techniques.

### 2.3 Spatio-Temporal Analysis

Scrub typhus-positive cases reported from May 2005 to April 2024 across all five study districts were used for the spatio-temporal analysis. The required block-wise shapefiles for the study districts were obtained from the Survey of India website (https://surveyofindia.gov.in/). The residential address of each confirmed scrub typhus case was used to determine the taluk of each patient for the analysis. The analysis was performed using QGIS software version 3.38.3. Block-wise shapefiles of all five districts were added to the software for spatial mapping. For the ease of understanding, six different maps were generated. The first map illustrated the district boundaries of all study districts. The second map showed the overall distribution of scrub typhus-positive cases over the 19-year study period. The third to fifth maps depicted the distribution of cases in five-year intervals starting from May 2005 to April 2020. The final map showed the distribution of cases over the most recent 4-year period, from May 2020 to April 2024. All spatial datasets, including block-level district shapefiles, were compiled and integrated with scrub typhus case data in QGIS according to the respective study periods.

### 2.4 Spatial Hotspot Analysis

The Positive Cases were used to perform a hotspot analysis (Getis-Ord Gi*) to identify the hot and cold spots for each pincodes of the five districts (Chittoor, Vellore, Ranipet, Tirupattur, and Tiruvannamalai). The hotspot analysis tool in ArcMap was used. Based on the positive cases of the pincodes, the spatial autocorrelation will work with the neighbouring pincodes. Getis-Ord Gi* statistics measure the spatial concentration between positive cases values and produce the Z-scores and p-values for each pincodes [23]. Whereas the Z-values are used to group pincodes into hot and cold spot categories, the P-value is used to differentiate them with confidence intervals (e.g., 90%, 95%, and 99%) for the hot and cold spots. The pincodes with dissimilar values from neighbouring pincodes is considered insignificant. The Getis-Ord G* statistics was performed through the formula mentioned below.

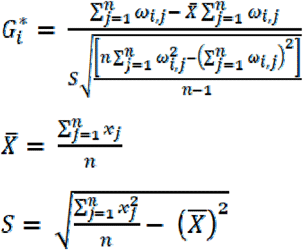

Where *x_j_* is the attribute value for the spatial feature *j*, *ω_i,j_* is the spatial weight between features *i* and *j, n* is the total number of features [23] [24].

The Gi* statistic returned for each feature in the dataset is a z-score. For statistically significant positive z-scores, the larger the z-score, the more intense the clustering of high values (hot spots) and negative z-scores, the smaller the z-score, the more intense the clustering of low values (cold spots).

### 2.5 Feature Engineering and Feature Selection

To capture the delayed effects of climate on scrub typhus transmission dynamics, lagged climatic features were created for each predictor, considering lag periods from 1 to 4 time steps. Also, rolling-window features such as moving averages and moving standard deviations were generated to quantify short-term climatic variability. Temporal indicators, including month, week-of-year, and seasonal encoding, were included to model seasonality. To reduce redundancy among features, multicollinearity was assessed using Pearson correlation and ExtraTrees regressor, and highly correlated features were removed based on predefined thresholds. To characterize the climatic and vegetation influence on disease incidence, a multivariate predictor set was constructed using environmental variables, including relative humidity, dew point temperature, evaporation, precipitation, atmospheric pressure, skin temperature, soil temperature, mean temperature, minimum temperature, maximum temperature, and NDVI area.

Let denote the observed disease case count at month, and let represent the value of the -th climatic predictor at the same month. Since climatic effects on disease transmission are delayed, lagged features were modelled. For each predictor, a lagged feature was computed as thereby allowing the models to learn short-term delayed associations between climate and disease incidence. To incorporate temporal structure and seasonality, month-of-year information was introduced through both categorical/ordinal and cyclic encodings. Specifically, with *m_t_* ∈ {1, … 12} denoting the month at time *t* cyclic seasonality was encoded as

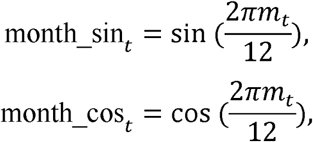

and a monotonic time index *I_t_* = *t* was included to represent long-term changes and nonstationarity.

To further separate systematic temporal components from irregular fluctuations, decomposition-based features were derived from the disease time series. In particular, classical decomposition and STL decomposition were employed to express the series as an additive model:

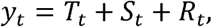

where *T_t_*, *S_t_* and *R_t_* denote the trend, seasonal, and residual components, respectively. These engineered features provided structured information on long-term progression and recurring seasonal burden, thereby improving the downstream models’ ability to capture non-linear temporal dynamics.

After feature construction, a two-stage selection strategy was adopted to retain informative features and reduce redundancy. First, an ExtraTrees regressor was fitted to estimate feature importance scores. Let *M* denote the trained ensemble model and *I_k_* denote the importance score assigned to feature *k.* The top *K* = 20 features were retained:

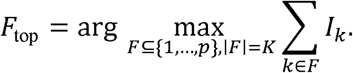

Second, multicollinearity among the selected features was controlled using correlation thresholding. Pearson correlation coefficients were computed for all feature pairs:

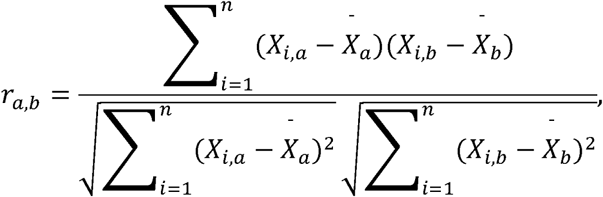

and an absolute correlation matrix |*r_a,b_*| was formed. A feature *b* was removed if it exhibited high correlation with any other feature *a*, based on a predefined threshold *τ* = 0.75

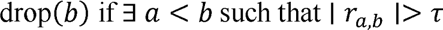

This procedure was implemented by examining only the upper-triangular portion of the correlation matrix to avoid duplicate comparisons. The final reduced feature set was then used for all models. The complete pipeline (lag generation, seasonal encoding, decomposition features, Extra-Trees ranking, and correlation pruning) was applied for each district to account for localized climatic characteristics and district-specific transmission dynamics. The finalized features retained for each district are summarized in Table 1 and were used throughout the modelling experiments.

**Table 1:** Final district-wise selected features used for time series modelling of Scrub Typhus incidence.

| **District** | **Final selected features (used for modelling)** |
| --- | --- |
| **Chittoor** | Residual component, STL seasonal component, NDVI area (lag1), STL trend, NDVI area percent, NDVI area (lag2), time index, pressure (lag3), NDVI area (lag3), month, precipitation (lag1), relative humidity (lag3), and soil temperature. |
| **Ranipet** | Residual component, STL seasonal component, trend component, NDVI area (lag2), NDVI area (lag1), time index, relative humidity (lag1), evaporation (lag1), NDVI area percent, precipitation (lag2), precipitation (lag3), NDVI area (lag3), pressure (lag3), and evaporation (lag3). |
| **Tirupattur** | Residual component, STL residual, STL seasonal component, STL trend, month, NDVI area (lag2), pressure (lag3), time index, precipitation, and precipitation (lag2). |
| **Tiruvannamalai** | Trend component, residual component, STL seasonal component, relative humidity (lag2), maximum temperature, time index, evaporation, seasonal component, and precipitation. |
| **Vellore** | Trend component, residual component, relative humidity (lag1), month cosine, STL seasonal component, NDVI area (lag3), precipitation, evaporation, time index, month, and seasonal component. |

### 2.6 Modelling frameworks

In this study, scrub typhus incidence forecasting was performed using three model types: classical time-series models, ML regression models, and DL sequence models. Classical statistical approaches include ARIMA [25], SARIMA [25] and exponential smoothing [26], which models temporal structure using univariate assumptions and relies on trend, seasonality, and autocorrelation patterns. In parallel, scrub typhus forecasting is formulated as a supervised regression task using ML models. Ridge regression [27] is applied to mitigate overfitting by L2-regularizing regression coefficients. ElasticNet regression [28] combines L1 and L2 penalties, enabling both coefficient shrinkage and implicit feature selection. Random forest regression [29] is used as an ensemble method that aggregates predictions from multiple decision trees trained on bootstrapped samples, improving the reduction of variance. Gradient boosting regressor [30] builds decision trees, where each successive tree corrects the residual errors of the previous ensemble. XGBoost [31] extends gradient boosting with efficient optimization, strong regularization, and scalable training, thereby improving predictive accuracy. LightGBM [32] further enhances efficiency through histogram-based splitting and leaf-wise tree growth, making it suitable for high-dimensional feature spaces. CatBoost [33] incorporates ordered boosting and built-in regularization strategies, which reduce prediction bias and overfitting, particularly in complex nonlinear relationships.

Also, several DL architectures are proposed to capture nonlinear and long-term temporal dependencies. A deep neural network [34] consists of multiple fully connected layers that learn complex nonlinear mappings between climate variables and disease incidence. A standard long Short-term memory (LSTM) [35] network leverages gated memory units to model temporal dependencies and handle vanishing gradient issues in sequential data. A stacked LSTM [35] extends this architecture by stacking multiple LSTM layers, enabling the learning of hierarchical temporal representations. Temporal convolutional networks (TCN) [36] employ one dimensional causal convolutions, ensuring that predictions at a given time step depend only on current and past inputs. Finally, a bidirectional long short-term memory (BiLSTM) [37] processes sequences in both forward and backward directions, integrating past and future contextual information within the input window to learn richer temporal patterns.

### 2.7 Model evaluation

To ensure an unbiased assessment of forecasting performance, all models were evaluated using a strict chronological split to prevent information leakage. The time series dataset was split into a training period from May 2005 to December 2021 and a test period from January 2022 to May 2024. Models were fitted on the training set, and predictions were generated for the test set using the trained parameters. Model performance was quantified using complementary error-based and goodness-of-fit metrics, including RMSE, and *R*^2^, which capture average deviation, sensitivity to large errors, and explained variance.

The RMSE emphasizes larger deviations and is computed as

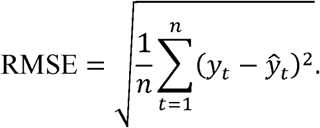

To evaluate the proportion of variance in observed scrub typhus cases explained by the model, *R*^2^ was calculated as

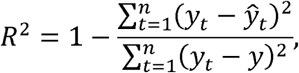

where *y* denotes the mean of observed values over the evaluation period. For consistency across districts and modelling paradigms.

### 2.8 Ethical Considerations

The study was approved by the Institutional Review Board of Christian Medical College, Vellore (IRB Approval No: 15919 dated 22.11.2023). As this was a retrospective analysis of anonymized medical records, the IRB granted a waiver of individual informed consent. All patient identifiers were removed prior to data analysis, and confidentiality was strictly maintained in accordance with the principles of the Declaration of Helsinki.

## 3 Results and Discussion

### 3.1 Characteristics of Scrub Typhus Cases

A total of five district-time observations were analysed over the period May 2005 to May 2024. Scrub typhus incidence showed substantial temporal variability, and across the study period, Vellore recorded the highest mean incidence. Table 2 presents the demographic characteristics of the scrub typhus cases analysed in this study.

**Table 2:** Demographic characteristics of Chittoor, Ranipet, Tirupattur, Vellore, and Tiruvannamalai, May 2005 to May 2024.

| Category | Group | Tested Numbers | Tested (%) | ST positives (Numbers) | Positive (%) |
| --- | --- | --- | --- | --- | --- |
| Gender | M | 10840 | 51.2 | 2519 | 46.2 |
| Gender | F | 10319 | 48.7 | 3127 | 55.4 |
| Age | 0-20 years | 2188 | 10.3 | 389 | 6.9 |
| Age | 21-30 years | 4596 | 21.7 | 970 | 17.2 |
| Age | 31-40 years | 3554 | 16.8 | 982 | 17.4 |
| Age | 41-50 years | 3503 | 16.5 | 1157 | 20.5 |
| Age | 51-60 years | 3479 | 16.4 | 1148 | 20.3 |
| Age | >60 years | 3852 | 18.2 | 1002 | 17.7 |
| Patient Type | IP | 11576 | 18.2 | 3213 | 56.9 |
| Patient Type | OP | 9596 | 45.3 | 2435 | 43.1 |
| **Total** | **All** | **21172** |  | **5648** |  |

### 3.2 Characteristics of Meteorological Factors

Table 3 summarizes the central tendency and variability of monthly scrub typhus cases and key meteorological variables across five districts from May 2005 to May 2024. It reports mean, dispersion, and percentile ranges, allowing you to assess typical conditions as well as extremes over the study period. Climatic variables exhibit substantial seasonal and interannual variability, particularly precipitation, evaporation, NDVI area, and temperatures, indicating pronounced hydroclimatic and vegetation dynamics in the region. Scrub typhus case counts exhibit right skewness, with a low median and a high maximum, reflecting sporadic outbreaks rather than uniform transmission. Overall, the statistics provide a quantitative baseline for examining associations among climate variability, environmental conditions, and scrub typhus incidence in the study districts.

**Table 3:** Summary statistics of monthly scrub typhus cases and associated meteorological variables in Chittoor, Ranipet, Tirupattur, Vellore, and Tiruvannamalai from May 2005 to May 2024.

| Variable | Total | Mean | SD | Min | P25 | Median | P75 | Max |
| --- | --- | --- | --- | --- | --- | --- | --- | --- |
| Dew Point (°C) | - | 18.9 | 2.0 | 11.2 | 18.0 | 19.3 | 20.4 | 22.8 |
| Pressure (hpa) | - | 967.8 | 11.5 | 948.6 | 957.7 | 966.7 | 975.6 | 991.6 |
| Skin Temperature (°C)* | - | 27.3 | 3.1 | 21.1 | 25.4 | 27.2 | 29.2 | 36.6 |
| Soil Temperature (°C) | - | 28.1 | 3.0 | 22.0 | 26.3 | 28.0 | 29.9 | 37.1 |
| Max. Temperature (°C) | - | 31.7 | 3.1 | 24.7 | 29.8 | 31.6 | 33.7 | 40.0 |
| Mean Temperature (°C) | - | 26.3 | 2.6 | 20.8 | 24.5 | 26.4 | 28.1 | 33.3 |
| Min. Temperature (°C) | - | 21.8 | 2.8 | 14.3 | 19.7 | 22.1 | 24.0 | 27.8 |
| wind_u_10m (m/s) | - | 0.41 | 1.6 | -1.95 | -1.03 | 0.0728 | 1.8 | 4.8 |
| wind_v_10m (m/s) | - | -0.05 | 0.65 | -1.90 | -0.38 | 0.01 | 0.3 | 2.1 |
| Evaporation (mm) | - | -594.4 | 832.7 | -3736.1 | -667.4 | -219.6 | -116.4 | -28.1 |
| Precipitation (mm) | - | 461.8 | 885.9 | 0.1 | 48.0 | 144.0 | 389.2 | 9068.1 |
| NDVI Area (%) | - | 26.3 | 26.3 | 0.0 | 3.2 | 17.5 | 46.9 | 95.0 |
| Relative Humidity (%) | - | 68.1 | 8.9 | 48.3 | 61.0 | 67.2 | 75.2 | 90.9 |
| Scrub Count Cases | 5648 | 5.0 | 8.1 | 0.0 | 0.0 | 2.0 | 6.0 | 73.0 |

\* temperature of the very top layer of the Earth's surface (land or water) or the top of a vegetative canopy

### 3.3 Climate variability and seasonal patterns of scrub typhus incidence

Figure 3 presents the long-term temporal covariation between monthly scrub typhus cases and multiple climatic and environmental variables over the study period. Scrub typhus exhibits strong seasonal peaks that recur annually and align closely with warmer temperatures, higher dew point, elevated relative humidity, and increased vegetation cover. Peaks in cases frequently follow periods of high precipitation and reduced evaporation, indicating favourable moist environmental conditions for vector survival and transmission. Temperature extremes show a clear annual cycle, with higher minimum and mean temperatures coinciding with increased case counts, while pressure displays an inverse seasonal pattern relative to disease incidence. NDVI area shows pronounced seasonal growth that temporally overlaps with higher case activity, suggesting a link between vegetation dynamics and disease ecology. Wind speed shows a limited direct association, whereas combined moisture-related variables demonstrate stronger coherence with outbreak periods. Overall, it highlights a consistent seasonal coupling between scrub typhus incidence and climate variability, supporting the role of temperature, moisture, and vegetation as key environmental drivers of disease transmission dynamics.

**Figure 3:**
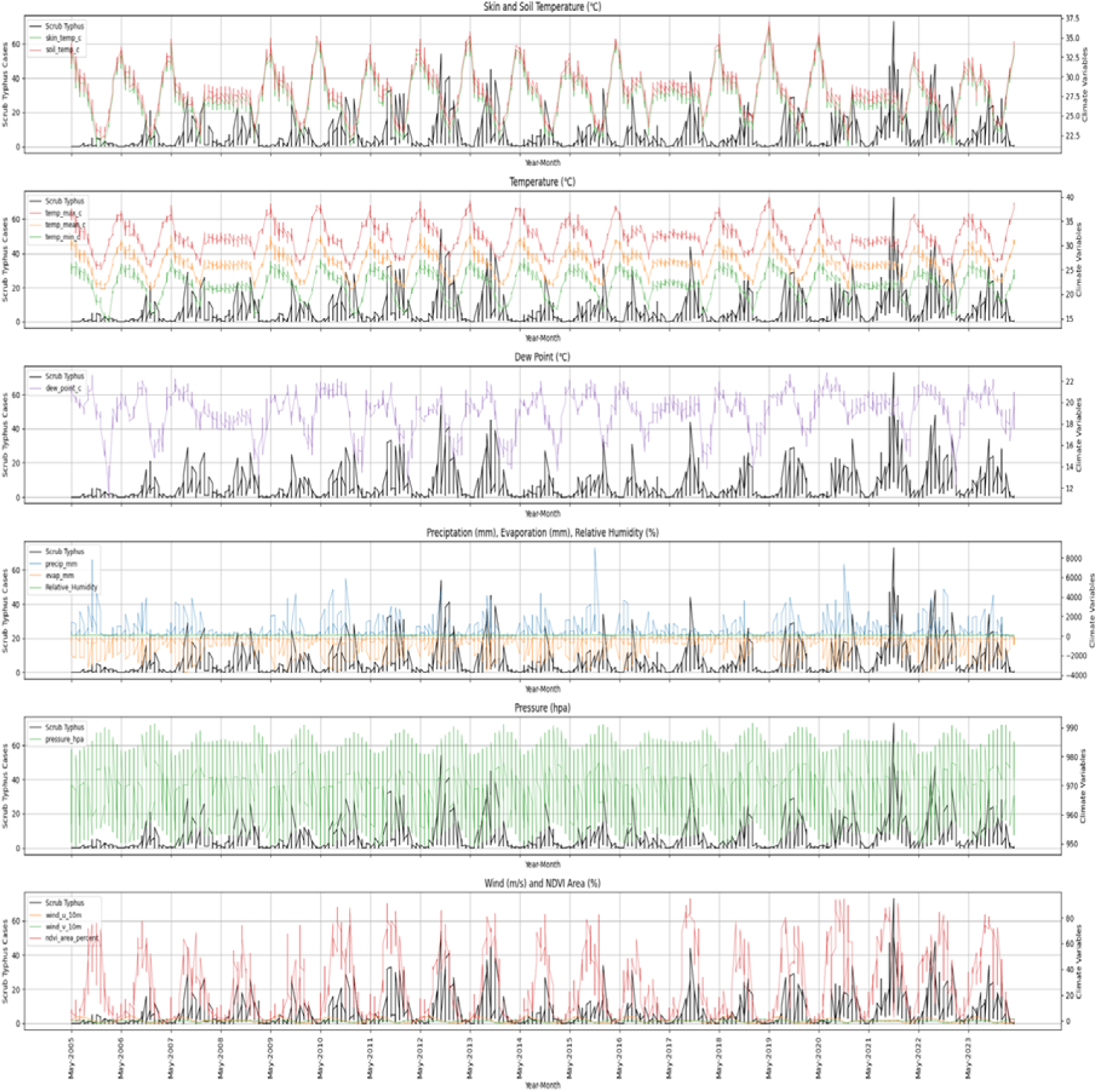
Illustrates the seasonal relationship between monthly scrub typhus cases and key climatic and environmental variables.

### 3.4 Spatio-Temporal Analysis

Figure 4 shows the taluk-wise distribution of scrub typhus cases in the districts of Vellore, Ranipet, Tirupattur, Chittoor, and Tiruvannamalai from May 2005 to April 2024. Figure 4a. illustrates the district boundaries and the location of CMC, Vellore. Figure 4b. presents the consolidated blockwise distribution of scrub typhus cases over the 19-year study period. Figures 4c. to 4f. shows the distribution of cases during four time intervals: May 2005–April 2010, May 2010–April 2015, May 2015–April 2020, and May 2020–April 2024.

**Figure 4:**
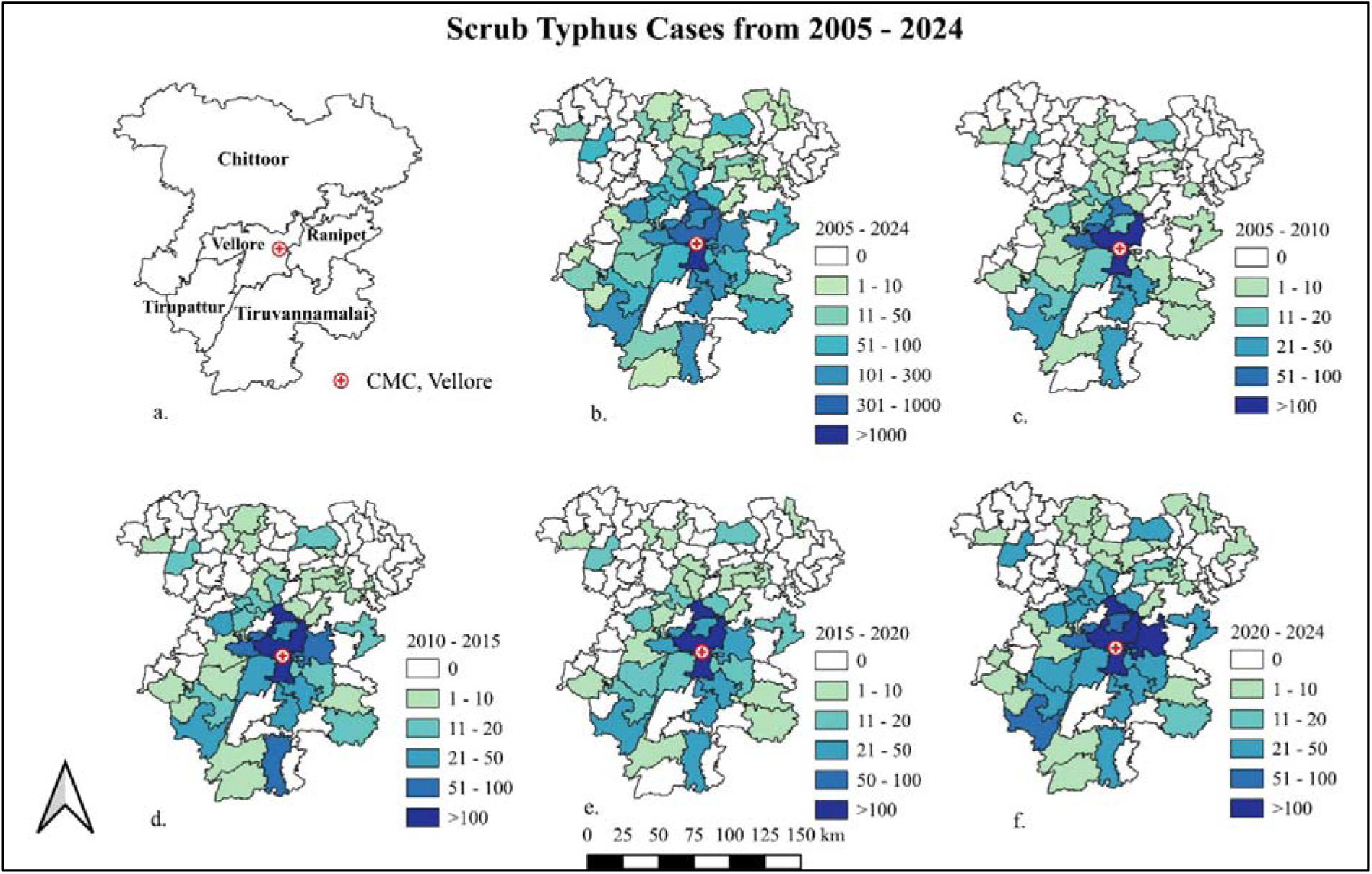
Taluk-wise scrub typhus cases from Vellore, Ranipet, Tirupattur, Tiruvannamalai & Chittoor districts from May 2005 to April 2024.

### Scrub typhus cases from May 2005 to April 2024

A total of 5,648 cases were reported from Vellore, Ranipet, Tirupattur, Chittoor and Tiruvannamalai districts from May 2005 to April 2024. Out of which, Vellore contributed 44.3% (2502) followed by Chittoor 32.5% (1833), Tiruvannamalai 11.2% (635), Ranipet 6.7% (381) and Tirupattur 5.3% (297). Sub-district analysis revealed that Vellore taluk in Vellore district (1,420 cases), Chittoor taluk in Chittoor district (955 cases), Tiruvannamalai taluk in Tiruvannamalai district (238 cases), Walaja Taluk of Ranipet district (244) and Triupattur taluk in Tirupattur district (181) were the major contributors to scrub typhus cases over the past 19 years. From May 2005 to April 2024, Vellore, Chittoor, and Katpadi taluks each reported more than 500 scrub typhus cases. Walaja, Tiruvannamalai, and Gudiyattam taluks reported 201–300 cases, whereas Tirupattur, Polur, Arni, Gudipala, and Palamaner taluks reported 101–200 cases.

### Scrub typhus cases from May 2005 to April 2010

A total of 900 cases from reported from Vellore, Ranipet, Tiruppatur, Chittoor and Tiruvannamalai districts from May 2005 to April 2010. Out of 900, Vellore contributed 437 (48.6%), Chittoor contributed 221 (24.6%), Ranipet 5.2% (47), Tirupattur 5.7% (51), and Tiruvannamalai district contributed 144 (16%) cases. Sub-district analysis revealed that Vellore taluk (236 cases), in Vellore district, Chittoor taluk (98 cases), in Chittoor district, Walaja Taluk in Ranipet district (30), Tirupattur Taluk of Tirupattur district (36) and Polur and Tiruvannamalai taluk (46 each) in Tiruvannamalai district were the major contributors to scrub typhus cases. From May 2005 to April 2010, Vellore taluk reported more than 200 cases, Katpadi taluk reported more than 100 cases, Chittoor and Gudiyattam taluks reported 50 – 100 scrub typhus cases.

### Scrub typhus cases from May 2010 to April 2015

A total of 1664 cases from reported from Vellore, Ranipet, Tirupattur, Chittoor and Tiruvannamalai districts from May 2010 to April 2015. Out of 1664, Vellore contributed 837 (50.3%), Ranipet 5.3% (89), Tirupattur district accounted for 4.4% (74), Chittoor contributed 446 (26.8%) and Tiruvannamalai district contributed 218 (13.1%) cases. Sub-district analysis revealed that Vellore taluk (476 cases), in Vellore district, Walaja Taluk (59 cases) in Ranipet campus, Tirupattur taluk (50 cases) of Tirupattur district, Chittoor taluk (232 cases), in Chittoor district and Tiruvannamalai taluk (100 each) in Tiruvannamalai district were the major contributors to scrub typhus cases. From May 2010 to April 2015, Vellore taluk reported more than 400 cases, Katpadi and Chittoor taluks reported more than 200 cases, Tiruvannamalai taluk reported 100 scrub typhus cases.

### Scrub typhus cases from May 2015 to April 2020

A total of 1207 cases were reported from Vellore, Ranipet, Tirupattur, Chittoor, and Tiruvannamalai districts from May 2015 to April 2020. Out of 1207, Vellore contributed 578 (47.9%), Ranipet contributed 63 (5.2%), Tirupattur contributed 52 cases (4.3%), Chittoor contributed 404 (33%), and Tiruvannamalai district contributed 110 (9%) cases. Sub-district analysis revealed that Vellore taluk (338 cases), in Vellore district, Walaja Taluk in Ranipet district (37 cases), Tirupattur Taluk (24 cases) in Tirupattur district, Chittoor taluk (230 cases), in Chittoor district, and Tiruvannamalai taluk (45 each) in Tiruvannamalai district were the major contributors to scrub typhus cases. From May 2015 to April 2020, Vellore taluk reported more than 300 cases, Chittoor taluk reported more than 200 cases, and Katpadi taluk reported more than 100 scrub typhus cases.

### Scrub typhus cases from May 2020 to April 2024

A total of 1877 cases were reported from Vellore, Ranipet, Tiruppattur, Chittoor, and Tiruvannamalai districts from May 2020 to April 2024. Out of 1877, Vellore contributed 650 (34.6%), Ranipet contributed 182 (9.7%), Tirupattur contributed 120 (6.4%), Chittoor contributed 762 (40.6%), and Tiruvannamalai district contributed 163 (8.7%) cases. Sub-district analysis revealed that Chittoor taluk (395 cases) in Chittoor district, Vellore taluk (370 cases), in Vellore district, Walaja Taluk (118 cases) in Ranipet district, Tirupattur taluk (71 cases) in Tirupattur district, Polur and Tiruvannamalai taluks (47 each) in Tiruvannamalai district were the major contributors to scrub typhus cases. From May 2020 to April 2024, Chittoor and Vellore taluks reported close to 400 cases, Katpadi taluk reported close to 200, and Walaja taluk reported 118 scrub typhus cases.

### 3.5 Spatial Hotspot Analysis

Figure 5 presents a hotspot analysis of positive scrub typhus cases using the Geits-ord GI*. The analysis identifies spatial clustering of reported positive cases across the study area. Each point represents an administrative unit, with circle size indicating the number of positive cases. Larger circles correspond to higher case counts. Hotspots shown in red indicate areas with significant clusters of high positive cases. Darker red indicates higher confidence, with 99 percent confidence indicating the strongest clustering. Coldspots shown in blue indicate significant clusters of low positive cases. Darker blue represents higher confidence coldspots. The central region around Vellore and nearby areas shows strong high-value hotspots at 95-99% confidence. This indicates intense spatial concentration of positive cases in this zone. Surrounding regions show a mix of moderate hotspots and non-significant areas, suggesting spatial spillover but at lower intensity. Peripheral areas, including Tirupattur, Ranipet, and Tiruvannamalai, show scattered cold spots and no significant patterns. These areas show lower-case clustering or a random spatial distribution. Overall, the map demonstrates clear spatial heterogeneity in positive cases. Central districts act as core transmission zones, while outer regions show weaker or insignificant clustering. This pattern highlights priority areas for targeted surveillance and intervention.

**Figure 5:**
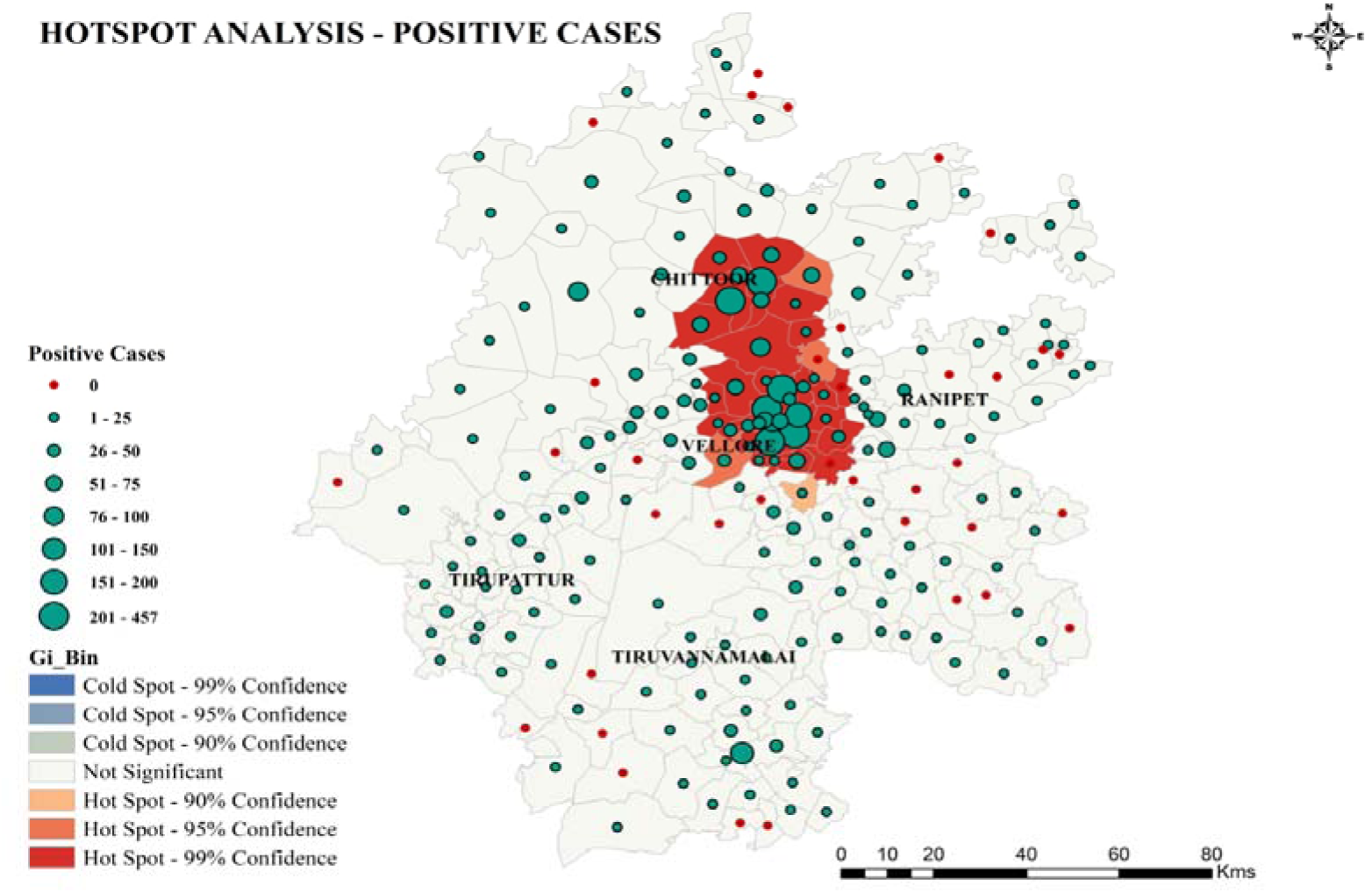
Getis-Ord Gi hotspot analysis of scrub typhus positive cases across Chittoor, Ranipet, Tirupattur, Vellore, and Tiruvannamalai districts (2005–2024). Significant hotspots (99% confidence, red) cluster in central Ranipet and Vellore district, with moderate hotspots (90–95% confidence, orange) in Tirupattur; coldspots (blue) appear peripherally. Case density gradients (1– 180+ cases) highlight transmission foci along Eastern Ghats scrub corridors.

### 3.6 Seasonality of Transmission

Scrub typhus cases displayed pronounced seasonality, revealing a clear and consistent seasonal pattern, with a marked increase in cases during the post-monsoon and early winter period, from August to December, and peak intensities are most observed in October and November, while the lowest case burden occurs from January to April (Figure 6). This seasonal rise is epidemiologically plausible because post-monsoon conditions, such as increased humidity, moderate temperature, and dense vegetation, promote the survival and proliferation of chigger mites and their rodent hosts, thereby increasing the probability of human exposure during intensified agricultural and outdoor activities in this period.

**Figure 6.**
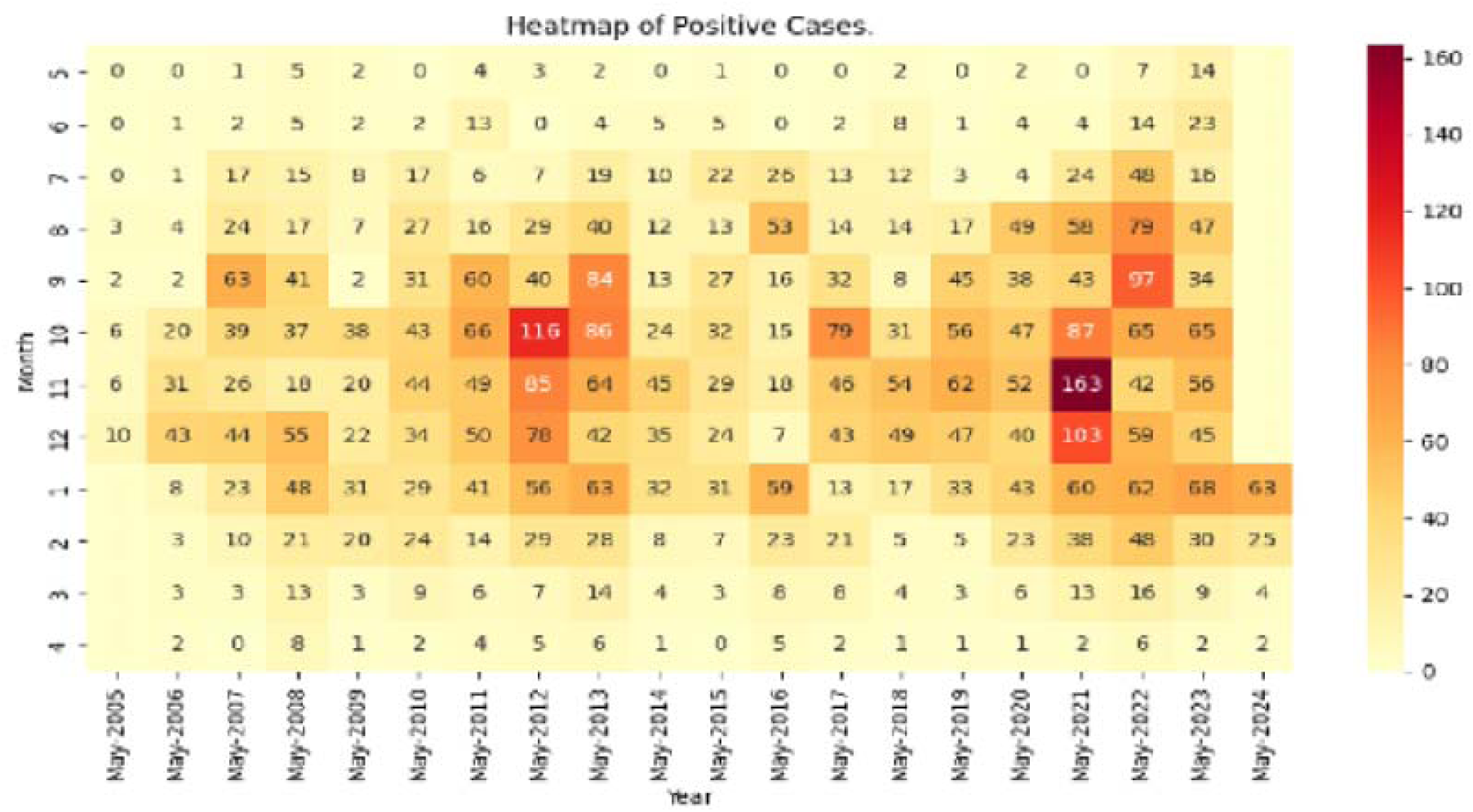
Heat Map showing the temporal seasonal impact of scrub typhus cases over the years from May 2005 to May 2024

The yearly trend analysis of scrub typhus-positive cases across the study districts shows substantial temporal variability, with distinct epidemic waves rather than a stable linear pattern (Figure 7). Overall, cases show an initial rise from 2005 onwards, followed by a pronounced peak during 2012–2013, a subsequent decline around 2014–2015, and then a renewed escalation culminating in the highest burden during 2021–2022, after which cases decrease again towards 2024. Across districts, Vellore and Chittoor account for the largest share of the case burden and drive the overall trend, indicating persistent higher transmission intensity in these districts compared to Ranipet, Tirupattur, and Tiruvannamalai, which exhibit lower but synchronized fluctuations.

**Figure 7.**
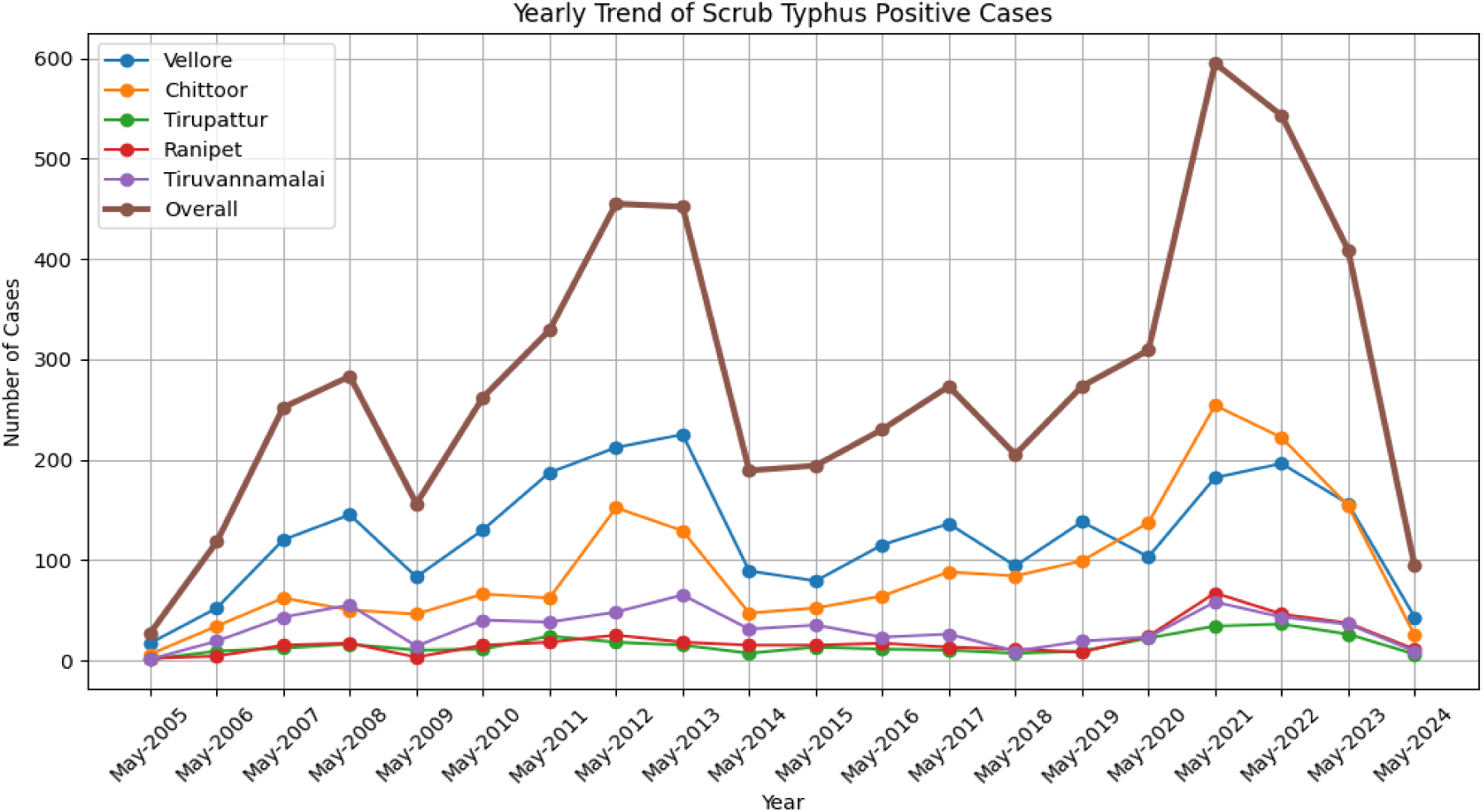
Yearly Trend of Scrub Positive cases of Chittoor, Ranipet, Tirupattur, Vellore, and Tiruvannamalai May 2005 to May 2024

The monthly trend of scrub typhus positive cases shows a strong and consistent seasonal cycle across all districts, with the lowest number of cases observed during the late spring and early summer period (April to June), followed by a steady increase from July onwards and a sharp rise during the post-monsoon season (Figure 8). The overall case burden peaks in October, remains high in November and December, and then declines during January to March, indicating that transmission intensity is highest during post-monsoon and early winter months. District-wise patterns are synchronized with Vellore and Chittoor contributing the greatest share of cases throughout the year and driving the overall seasonal peak, while Tirupattur, Ranipet, and Tiruvannamalai show lower magnitudes but similar temporal trends. This seasonal amplification is consistent with ecological conditions after monsoon rainfall, including increased humidity and vegetation cover that enhance chigger survival and rodent host activity, as well as increased human exposure from agricultural and outdoor activities during this period.

**Figure 8.**
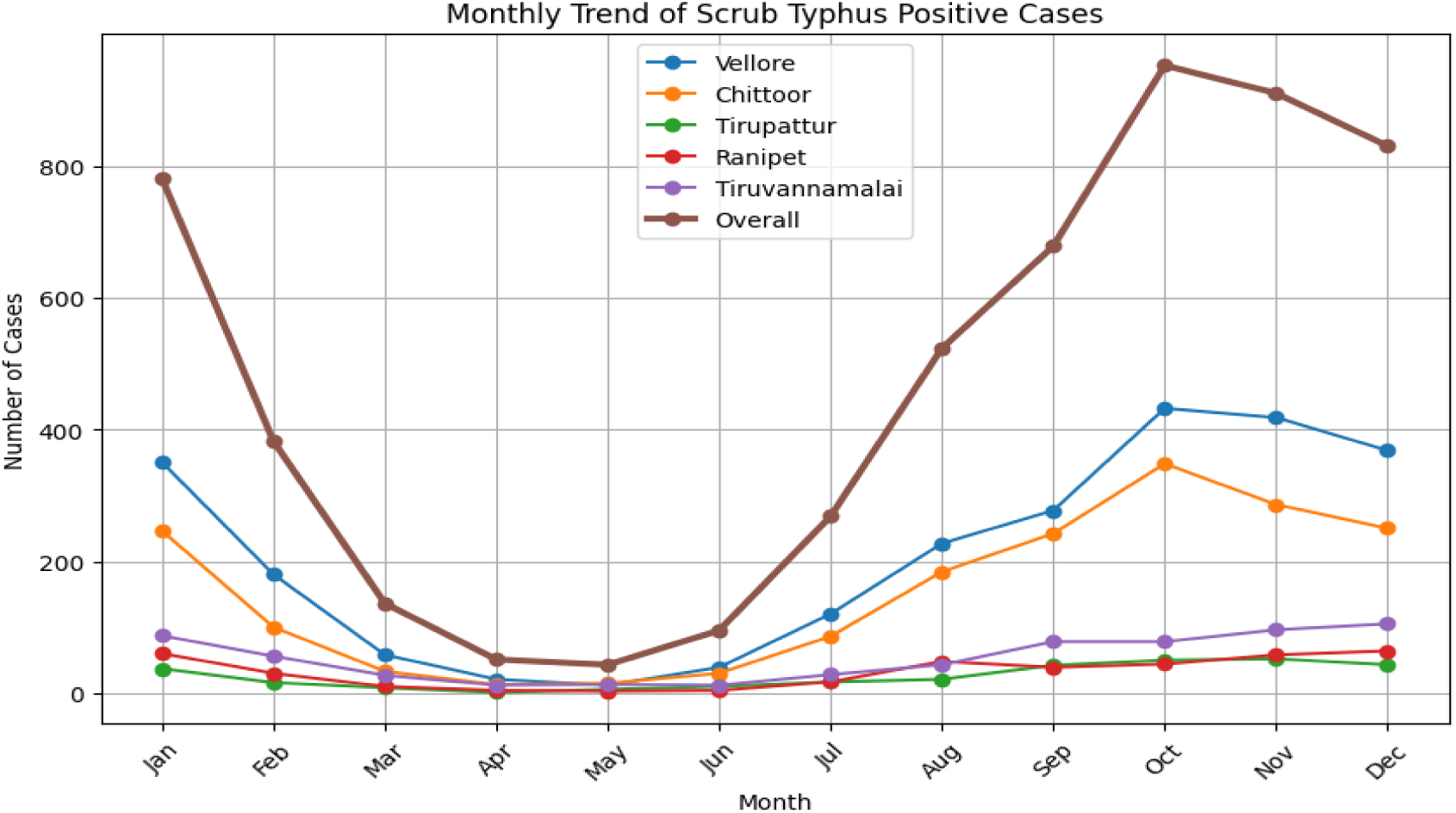
Monthly Trend of Scrub Positive cases of Chittoor, Ranipet, Tirupattur, Vellore, and Tiruvannamalai May 2005 to May 2024

The joinpoint regression plot of monthly scrub typhus cases indicates a statistically meaningful change in the long-term trend around 2019, as highlighted by the identified joinpoint (Figure 9). Prior to this time point, monthly case counts remain low, with sporadic fluctuations and limited growth; however, after 2019, the fitted regression line shows an upward trend, with more frequent and larger monthly peaks. The observed data points also demonstrate greater dispersion in the later years, reflecting stronger inter-month variability and more intense outbreak months. Overall, this pattern suggests a transition to a higher transmission phase in the post-2019 period, driven by changing environmental suitability, vector and reservoir dynamics, increased human exposure, and/or improved surveillance and reporting, resulting in a sustained rise in monthly scrub typhus incidence.

**Figure 9.**
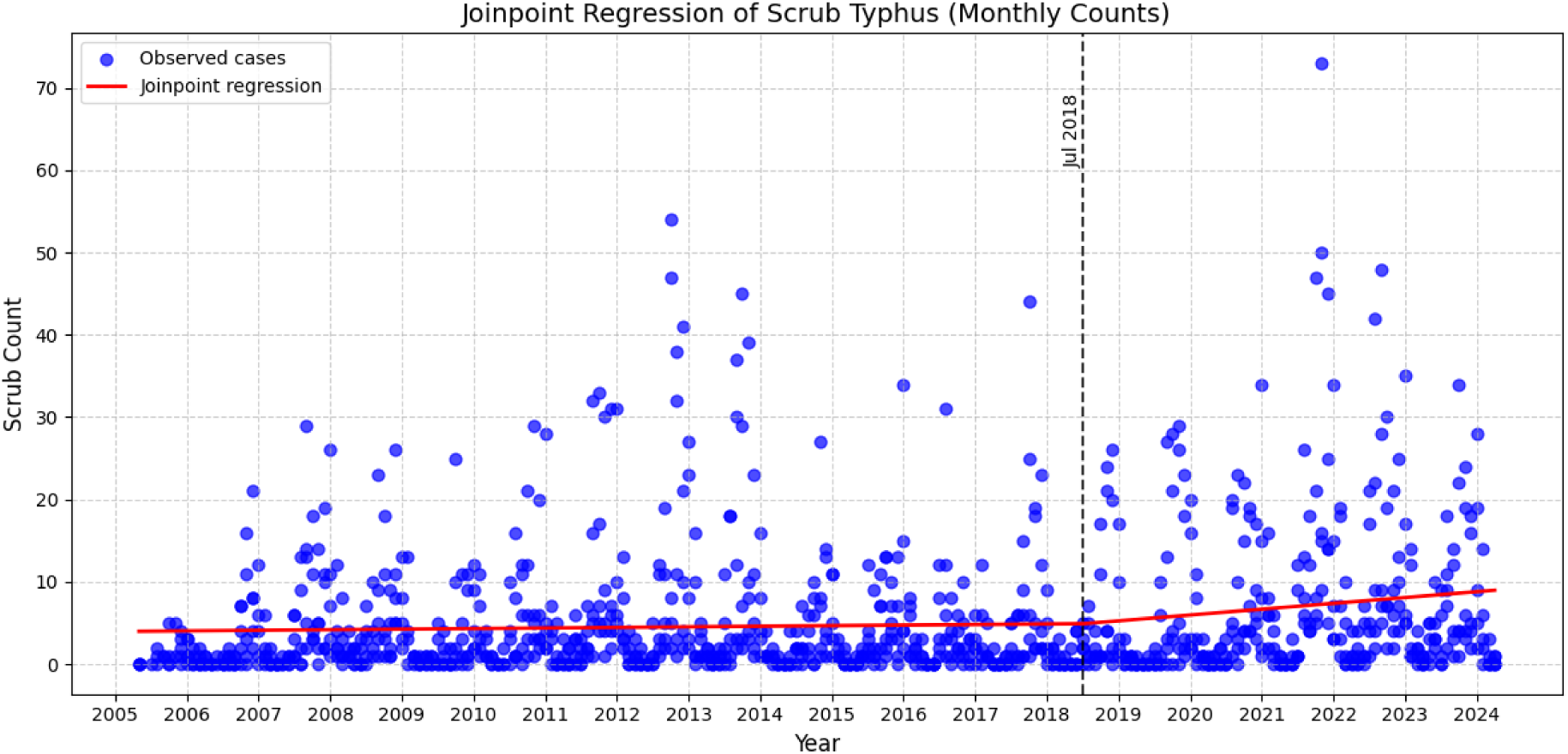
Joinpoint regression analysis of scrub typhus cases in Chittoor, Ranipet, Tirupattur, Vellore, and Tiruvannamalai, May 2005 to May 2024. The joinpoint indicates a statistically significant change in trend, and the regression lines show the annual percent change (APC) in each segment.

### 3.7 Scrub Typhus Cases Forecast Model Performance

Statistical baseline models provided reasonable short-term forecasting performance but were limited in their ability to capture nonlinear climate interactions. ML models achieved improved accuracy over statistical baselines across all districts. DL architectures further improved performance in districts exhibiting nonlinear temporal dynamics and abrupt outbreaks.

Table 4 presents model performance for the Chittoor district using RMSE and. Statistical models show weak performance. ARIMA and SARIMA record high RMSE and negative, indicating poor fit. Exponential Smoothing performs better, with lower RMSE and a positive, but remains inferior to learning based models. ML models show improved accuracy. RMSE decreases from LightGBM to Random Forest, while increases from near zero to 0.66. Random Forest, ElasticNet, and Ridge provide balanced performance with moderate error and variance explanation. DL models show mixed results. LSTM-based models perform poorly with high RMSE and negative or low. TCN shows great improvement, with lower RMSE and higher. DNN achieves the best performance overall, with the lowest RMSE and highest. These results indicate that DNN and TCN outperform other models for the Chittoor district.

**Table 4:** Performance metrics comparison table for the district of Chittoor.

| Model Category | Model | RMSE | $R^2$ |
| --- | --- | --- | --- |
| Statistical | ARIMA | 13.9163 | -1.4429 |
|  | SARIMA | 11.8314 | -0.7657 |
|  | Exp Smoothing | 5.9119 | 0.5591 |
| ML | LightGBM | 9.1164 | -0.0483 |
|  | Gradient Boosting | 8.5046 | 0.0876 |
|  | XGBoost | 8.0730 | 0.1779 |
|  | CatBoost | 5.9477 | 0.5538 |
|  | Ridge | 5.4641 | 0.6234 |
|  | ElasticNet | 5.4370 | 0.6271 |
|  | Random Forest | 5.1848 | 0.6609 |
| DL | LSTM | 12.1097 | -0.8498 |
|  | BiLSTM | 10.2313 | -0.3204 |
|  | Stacked LSTM | 8.7549 | 0.0332 |
|  | TCN | 4.7941 | 0.7101 |
|  | **DNN** | **3.7514** | **0.8225** |

Table 5 presents model performance for Ranipet district, measured by RMSE and *R*^2^. Statistical models show limited predictive ability. ARIMA reports negative *R*^2^ with high RMSE. Exponential Smoothing and SARIMA show slight improvement but retain low explanatory power. ML models show clear gains. RMSE declines from Random Forest to CatBoost. *R*^2^ increases from 0.30 to 0.94. CatBoost and Ridge achieve low error and high variance explanation. Dl models show gradual improvement with model complexity. LSTM performs weakly. BiLSTM and Stacked LSTM improve accuracy. TCN achieves low RMSE and high *R*^2^, comparable to the best ML models. These results show that CatBoost and TCN provide the most accurate forecasts for the Ranipet district.

**Table 5:** Performance metrics comparison table for the district Ranipet.

| Model Category | Model | RMSE | $R^2$ |
| --- | --- | --- | --- |
| Statistical | ARIMA | 2.9645 | -0.2335 |
|  | Exp Smoothing | 2.6313 | 0.0282 |
|  | SARIMA | 2.6000 | 0.0512 |
| ML | Random Forest | 2.2259 | 0.3046 |
|  | Gradient Boosting | 1.3603 | 0.7403 |
|  | LightGBM | 1.1728 | 0.8069 |
|  | ElasticNet | 0.7834 | 0.9139 |
|  | XGBoost | 0.7661 | 0.9176 |
|  | Ridge | 0.7313 | 0.9249 |
|  | **CatBoost** | **0.6645** | **0.9380** |
| DL | LSTM | 2.5029 | 0.1208 |
|  | BiLSTM | 1.9130 | 0.4864 |
|  | Stacked LSTM | 1.5938 | 0.6435 |
|  | DNN | 1.3195 | 0.7556 |
|  | TCN | 0.6693 | 0.9371 |

Table 6 presents model performance for Tirupattur district using RMSE and *R*^2^. Statistical models show weak performance. ARIMA, Exponential Smoothing, and SARIMA record high RMSE and negative *R*^2^, indicating poor model fit. ML models show improved accuracy. RMSE decreases from Random Forest to Ridge, while *R*^2^ increases from 0.32 to 0.76. Ridge and ElasticNet provide the best performance within this group. DL models show mixed results. LSTM performs poorly with negative *R*^2^. BiLSTM and Stacked LSTM show moderate improvement. TCN achieves the lowest RMSE and highest *R*^2^, indicating strong predictive performance. These results show that TCN outperforms other models for the Tirupattur district.

**Table 6:** Performance metrics comparison table for the district Tirupattur.

| Model Category | Model | RMSE | $R^2$ |
| --- | --- | --- | --- |
| Statistical | ARIMA | 3.0744 | -1.6074 |
|  | Exp Smoothing | 2.4518 | -0.6583 |
|  | SARIMA | 2.3308 | -0.4987 |
| ML | Random Forest | 1.5700 | 0.3200 |
|  | Gradient Boosting | 1.1296 | 0.6480 |
|  | LightGBM | 1.1077 | 0.6615 |
|  | XGBoost | 1.0518 | 0.6948 |
|  | CatBoost | 1.0451 | 0.6987 |
|  | ElasticNet | 0.9999 | 0.7242 |
|  | Ridge | 0.9335 | 0.7596 |
| DL | LSTM | 1.9760 | -0.0772 |
|  | BiLSTM | 1.5356 | 0.3495 |
|  | Stacked LSTM | 1.1480 | 0.6364 |
|  | DNN | 1.1171 | 0.6558 |
|  | **TCN** | **0.7345** | **0.8512** |

Table 7 presents model performance for the Tiruvannamalai district using RMSE and *R*^2^. Statistical models show very poor performance. Exponential Smoothing and ARIMA report high RMSE and strongly negative *R*^2^. SARIMA improves error levels but retains negative *R*^2^, indicating limited explanatory capacity. ML models perform strongly. RMSE decreases across models, while *R*^2^ increases from 0.80 to 0.99. Ridge, CatBoost, and LightGBM achieve low error and high variance explanation. DL models show high accuracy. LSTM provides moderate performance. BiLSTM, TCN, and Stacked LSTM achieve low RMSE and high *R*^2^. Stacked LSTM approaches the best ML results. These results indicate that ML models and advanced DL architectures outperform statistical methods for the Tiruvannamalai district.

**Table 7:** Performance metrics comparison table for the district Tiruvannamalai.

| Model Category | Model | RMSE | $R^2$ |
| --- | --- | --- | --- |
| Statistical | Exp Smoothing | 7.8845 | -7.8462 |
|  | ARIMA | 7.6608 | -7.3513 |
|  | SARIMA | 3.1331 | -0.3968 |
| ML | Random Forest | 1.1919 | 0.7978 |
|  | ElasticNet | 0.7482 | 0.9203 |
|  | XGBoost | 0.5523 | 0.9566 |
|  | Gradient Boosting | 0.5510 | 0.9568 |
|  | LightGBM | 0.3900 | 0.9784 |
|  | CatBoost | 0.3683 | 0.9807 |
|  | **Ridge** | **0.2919** | **0.9879** |
| DL | LSTM | 0.7917 | 0.9108 |
|  | DNN | 0.5917 | 0.9502 |
|  | BiLSTM | 0.4984 | 0.9646 |
|  | TCN | 0.4628 | 0.9695 |
|  | Stacked LSTM | 0.3504 | 0.9825 |

Table 8 presents model performance for the Vellore district using RMSE and *R*^2^. Statistical models show very poor performance. SARIMA, Exponential Smoothing, and ARIMA report high RMSE and negative *R*^2^, indicating weak model fit. ML models achieve high predictive accuracy. RMSE decreases across models, while *R*^2^ increases from 0.95 to above 0.99. Ridge, LightGBM, and CatBoost show low error and strong variance explanation. DL models perform well. A basic LSTM achieves lower accuracy than other approaches. BiLSTM, DNN, and TCN achieve low RMSE and high *R*^2^, approaching the best ML results. These results indicate that ML and selected DL models outperform statistical methods for the Vellore district.

**Table 8:** Performance metrics comparison table for the district of Vellore.

| Model Category | Model | RMSE | $R^2$ |
| --- | --- | --- | --- |
| Statistical | SARIMA | 17.6236 | -1.9476 |
|  | Exp Smoothing | 17.4208 | -1.8802 |
|  | ARIMA | 16.2231 | -1.4977 |
| ML | ElasticNet | 2.3537 | 0.9474 |
|  | Random Forest | 2.3172 | 0.9490 |
|  | Gradient Boosting | 1.5996 | 0.9757 |
|  | XGBoost | 1.0770 | 0.9890 |
|  | Ridge | 0.6586 | 0.9959 |
|  | LightGBM | 0.6341 | 0.9962 |
|  | **CatBoost** | **0.6303** | **0.9962** |
| DL | LSTM | 3.1401 | 0.9064 |
|  | Stacked LSTM | 2.2540 | 0.9518 |
|  | BiLSTM | 1.7605 | 0.9706 |
|  | DNN | 1.1906 | 0.9865 |
|  | TCN | 0.7624 | 0.9945 |

### 3.8 Forecast Visualization

Figure 10 presents district-level time-series forecasting results for monthly scrub typhus cases, using the best-performing model for each district. The panels correspond to Chittoor, Ranipet, Tiruvannamalai, Tirupattur, and Vellore. For Chittoor, the DNN model captures the long term trend and seasonal variability well, as reflected by an RMSE of 3.75 and *R*^2^ of 0.82. The historical series shows pronounced interannual variability with sharp epidemic peaks after 2020. The forecast follows the declining pattern after recent peaks but smooths extreme values, indicating limited sensitivity to sudden outbreak spikes. For Ranipet, CatBoost achieves a strong predictive performance with an RMSE of 0.66 and *R*^2^ of 0.94. The observed series exhibits low baseline incidence and occasional abrupt spikes. The CatBoost forecast aligns with recent observations and preserves short-term fluctuations that tree-based ensemble learning captures, nonlinear relationships, and abrupt changes in disease incidence. For Tiruvannamalai, Ridge regression yields near perfect performance with an RMSE of 0.29 and *R*^2^ of 0.99. The time series exhibits moderate variability with intermittent peaks. The forecast tracks observed values during the prediction window, indicating a stable linear relationship between predictors and scrub typhus incidence in this district. For Tirupattur, the TCN model provides improved performance compared to recurrent architectures, with an RMSE of 0.73 and of 0.85. The TCN forecast captures short-term temporal dependencies and maintains continuity with recent observations, though peak magnitudes are underestimated. For Vellore, CatBoost again demonstrates superior performance, with RMSE of 0.63. The district exhibits strong seasonality and recurrent high magnitude outbreaks. The forecast reproduces both the timing and relative intensities of recent peaks, highlighting the model’s ability to learn complex, nonlinear, and seasonal patterns. Overall, the figure demonstrates clear spatial heterogeneity in scrub typhus dynamics across districts and highlights that no single model is optimal. DL models perform better in districts with complex temporal dependencies, while ML models, such as CatBoost and Ridge, achieve superior accuracy in districts with stable or feature-driven patterns. These results give importance of district-specific model selection for reliable disease forecasting and public health planning.

**Figure 10:**
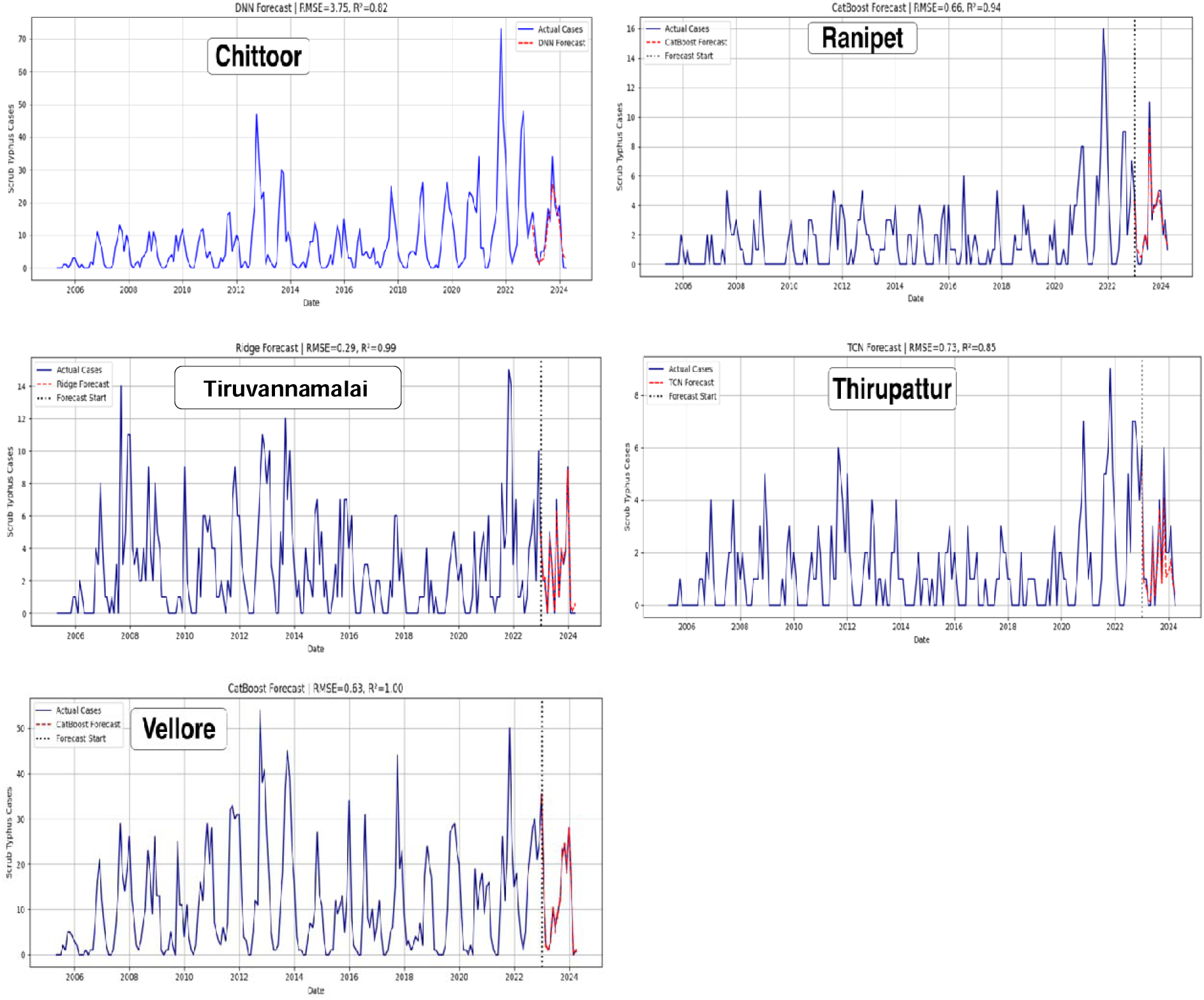
Scrub typhus fitting and forecasting results by the best proposed model for the five districts: Chittoor, Ranipet, Tirupattur, Vellore, and Tiruvannamalai. The vertical dash line separated the model fitting and forecasting parts.

## Conclusion

Scrub typhus transmission is strongly influenced by spatial heterogeneity, seasonal patterns, and climatic variability. Persistent seasonal peaks and clear climate associations highlighted the role of humidity, precipitation, vegetation, and temperature in shaping disease risk. Spatial analysis identified Vellore and Chittoor as stable core transmission zones, emphasizing the need for geographically targeted surveillance. Forecasting results showed that disease dynamics vary across districts, with no single model performing best in all locations. This supports the use of district-specific prediction strategies. Overall, the integrated spatiotemporal and forecasting framework provides a practical basis for early warning, focused interventions, and improved scrub typhus control.

## Data Availability

All data produced in the present study are available upon reasonable request to the authors

## Acknowledgments

The authors gratefully acknowledge the Indian Council of Medical Research (ICMR), New Delhi, India, for the research grants (EM/dev/SG/106/1363/2023 and DDR/CAREP-2023/0285). We acknowledge the help of Mrs. Ida Reena Rose, Ms. Ramya Krishnan and Dr. Aarush Sehgal in data entry.

## Supplementary Material

**Figure 1:**
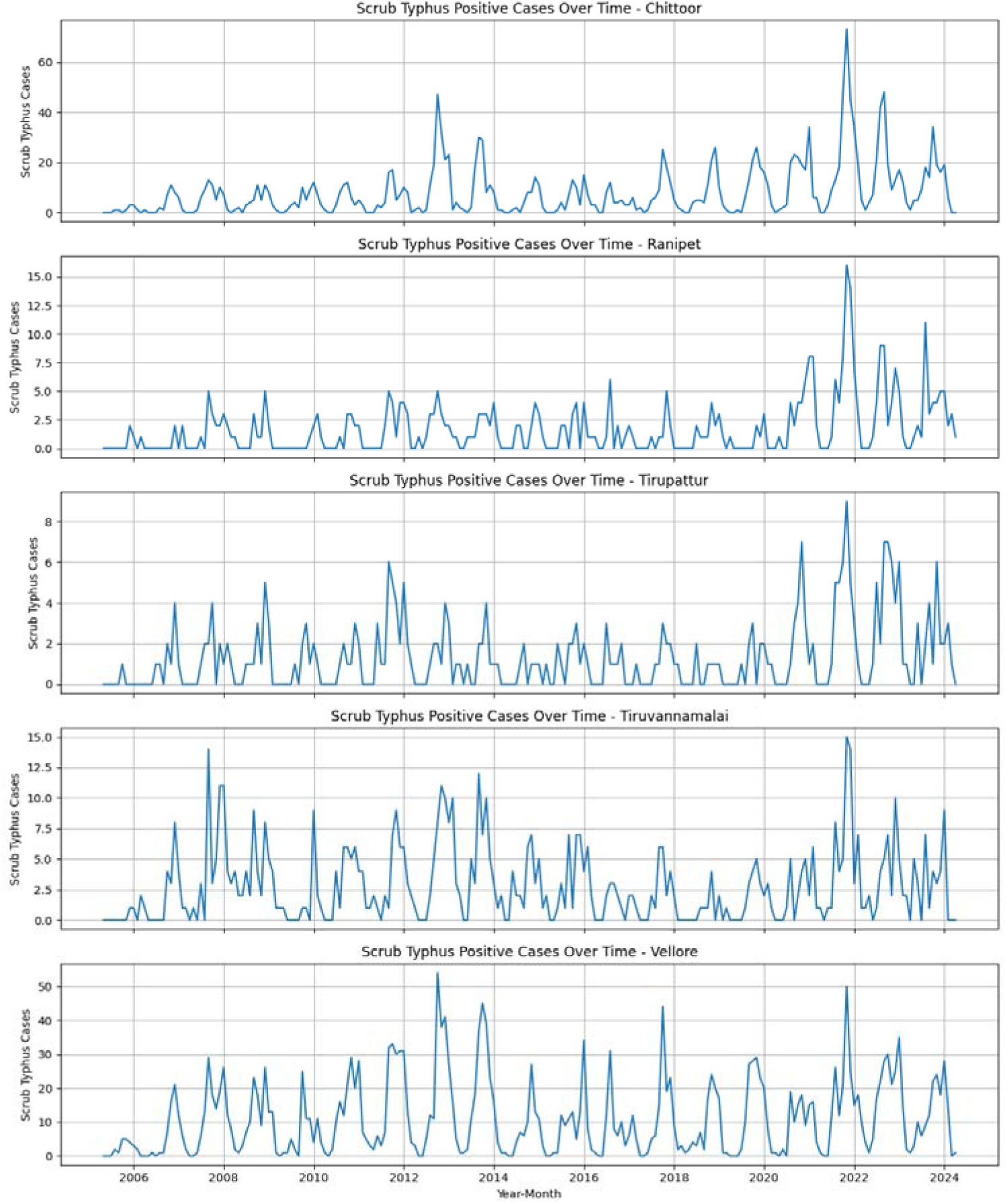
Illustrates the temporal variation in monthly scrub typhus positive cases across five districts, Vellore, Tiruvannamalai, Chittoor, Tirupattur, and Ranipet, from May 2005 to May 2024.

**Figure 2:**
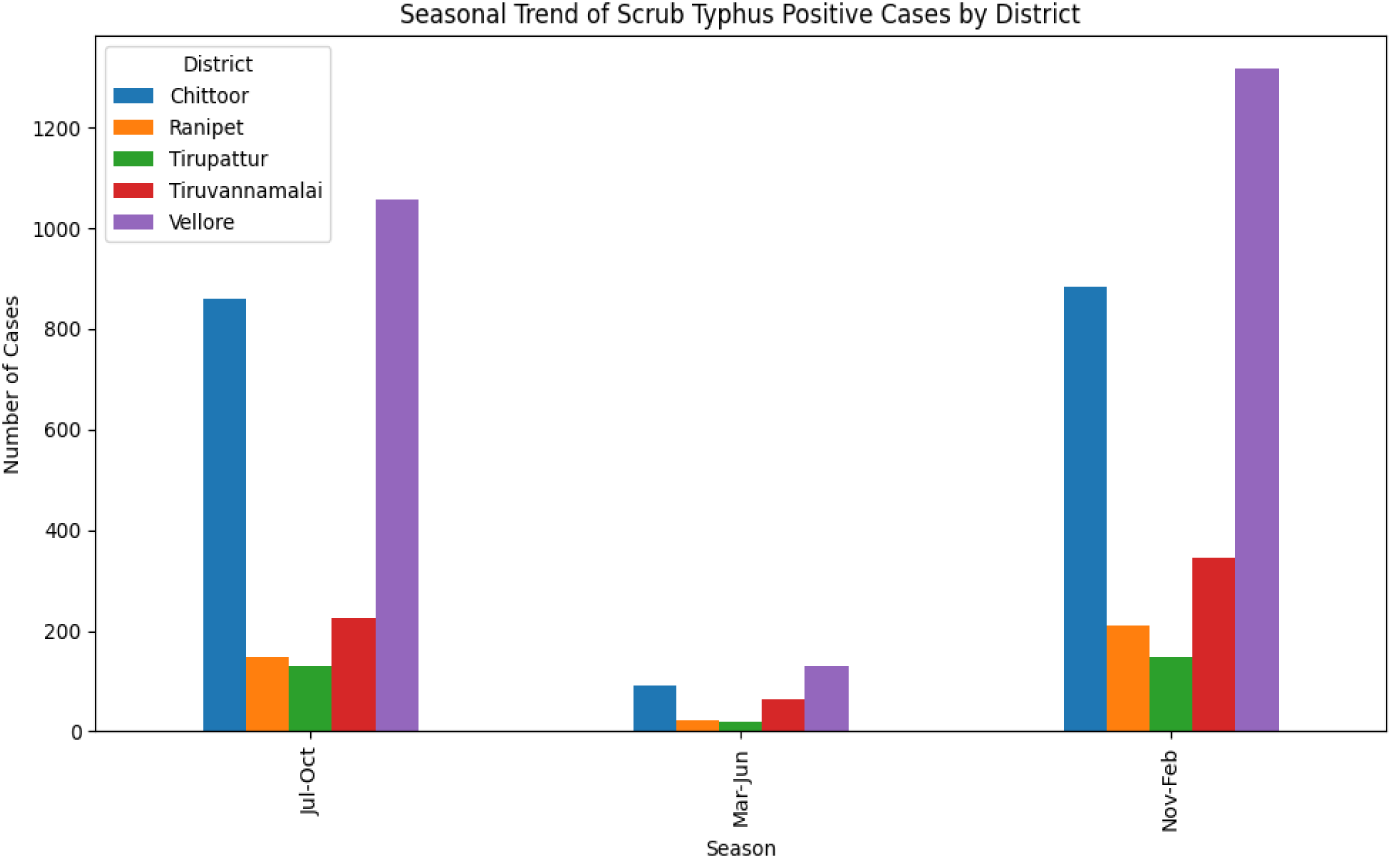
Seasonal Trend of scrub typhus Positive cases of Chittoor, Ranipet, Tirupattur, Vellore, and Tiruvannamalai for the duration May 2005 to May 2024

**Figure 3:**
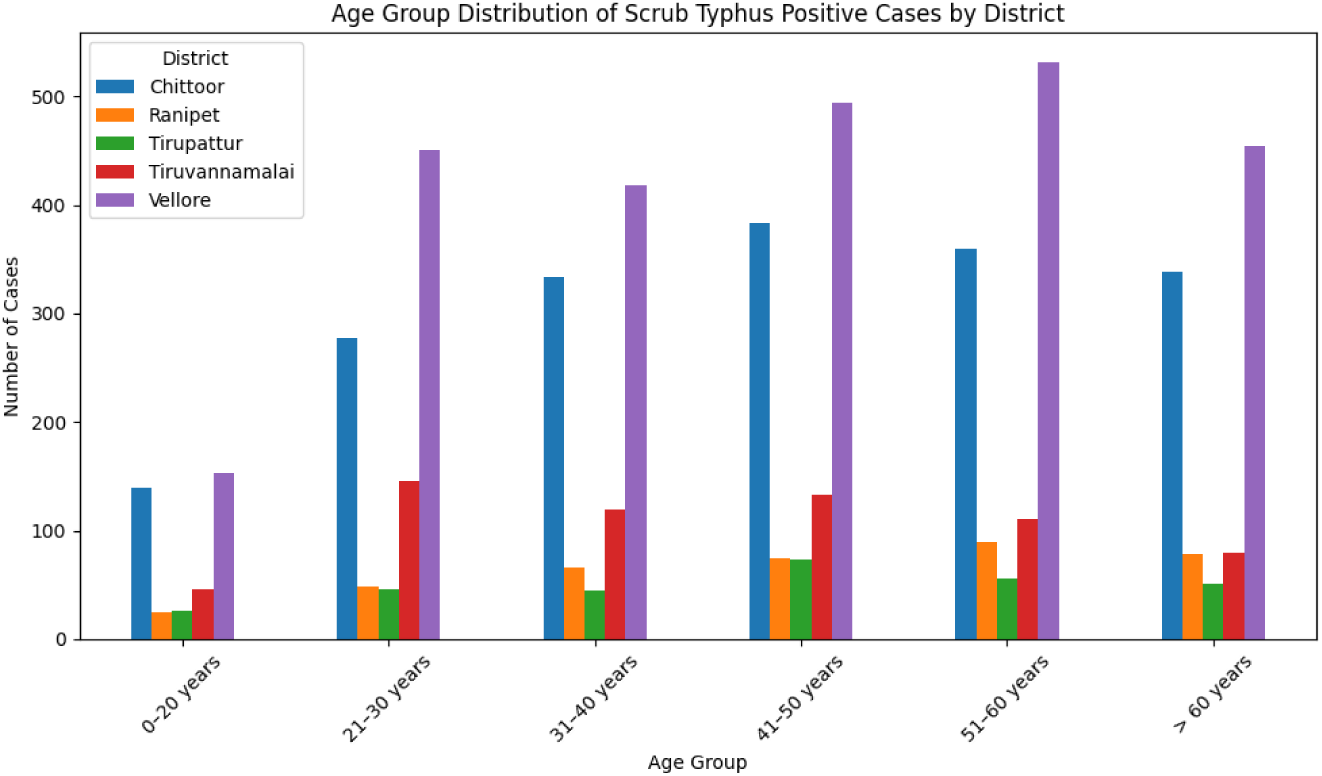
Age-wise Trend of scrub typhus Positive cases of Chittoor, Ranipet, Tirupattur, Vellore, and Tiruvannamalai for the duration May 2005 to May 2024

**Figure 4:**
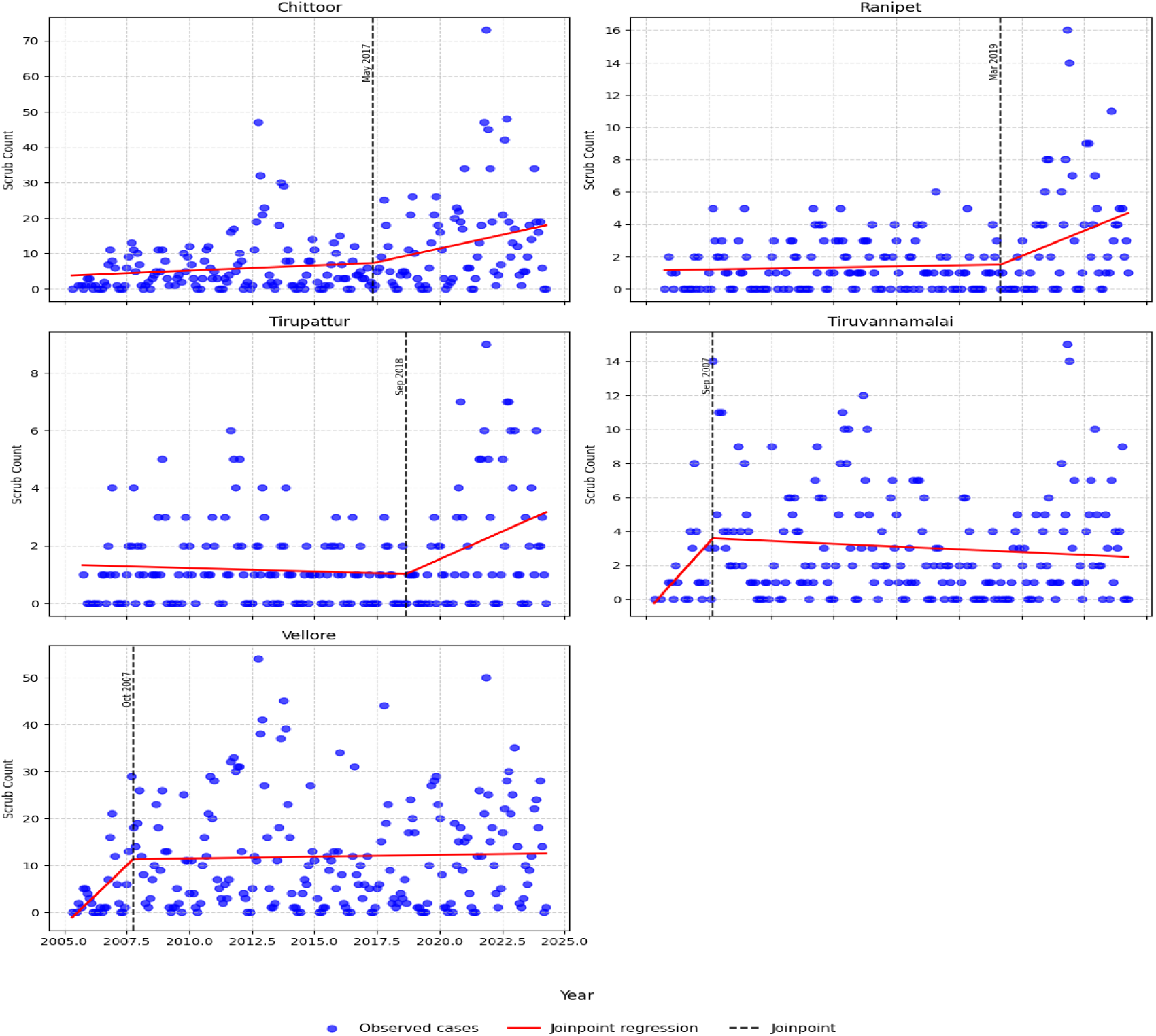
Joinpoint regression analysis of scrub typhus case trends in Chittoor, Ranipet, Tirupattur, Vellore, and Tiruvannamalai for the duration May 2005 to May 2024

**Figure 5:**
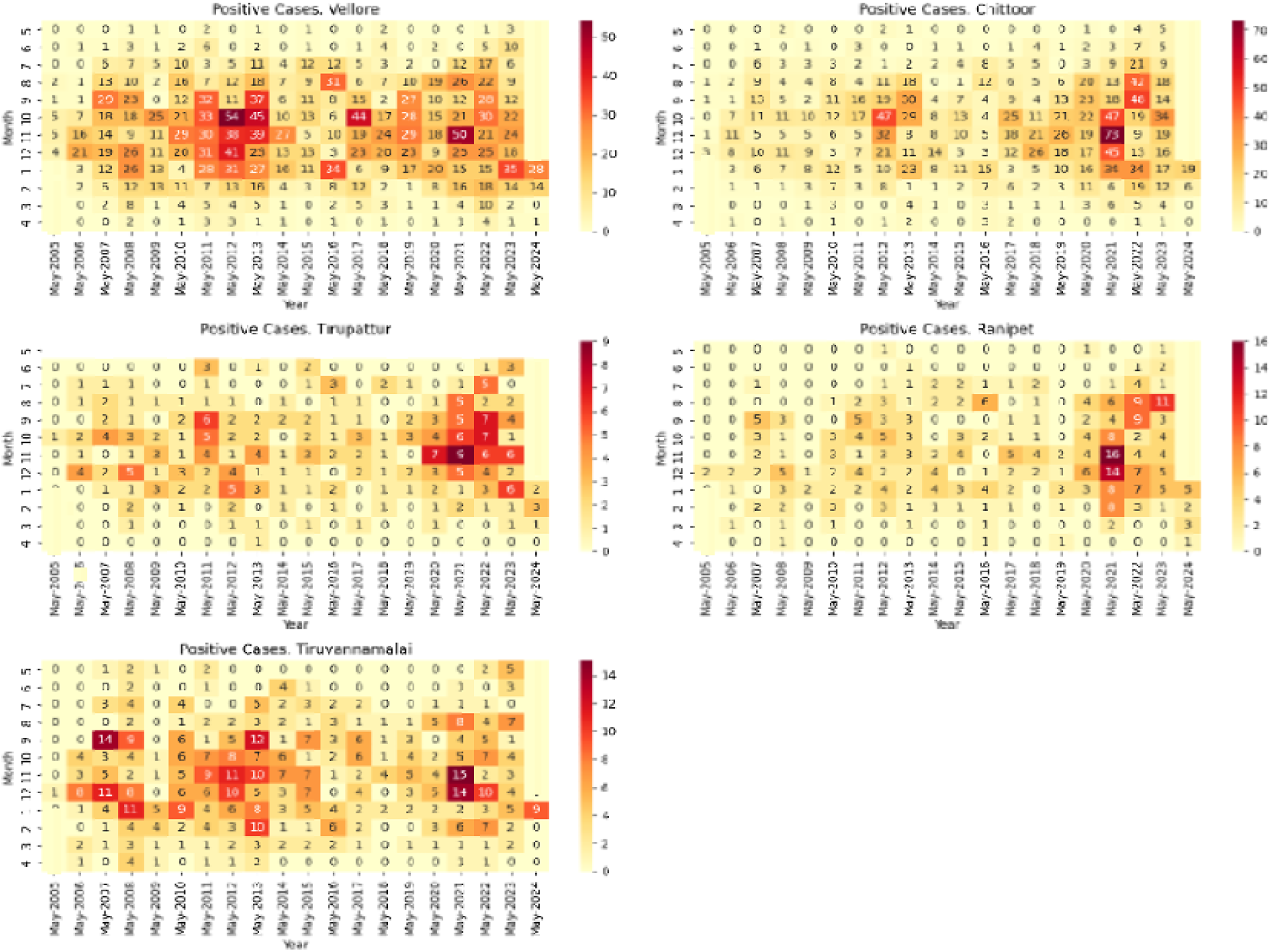
Heat map of scrub typhus case trends in Chittoor, Ranipet, Tirupattur, Vellore and Tiruvannamalai for the duration May 2005 to May 2024

**Figure 6:**
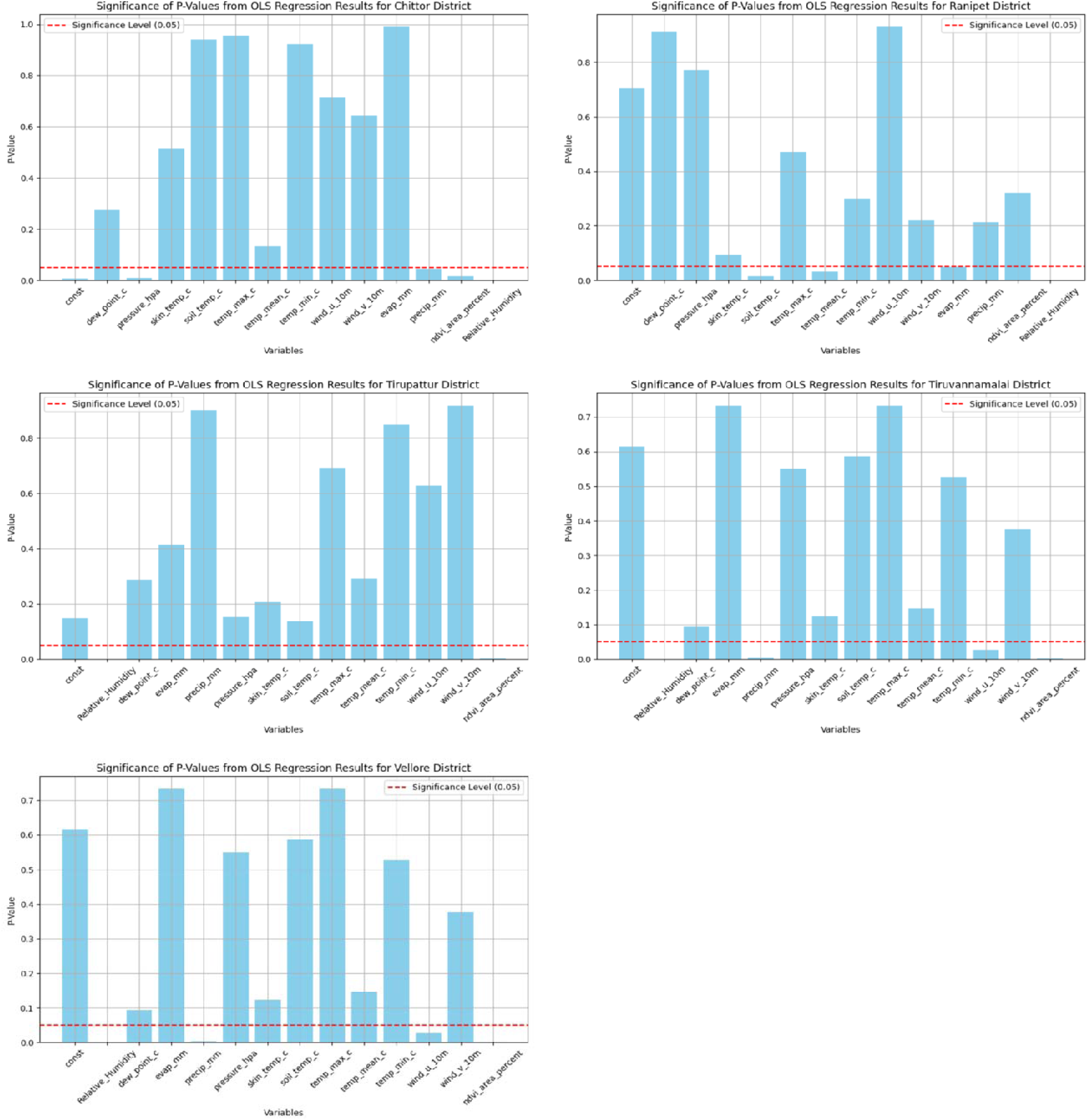
Significance of p-values from OLS regression analysis of scrub typhus risk factors across five study districts (May 2005–May 2024). Age group (21-30 years) (p<0.01) and occupation (others) (p<0.05) emerge as highly significant predictors.

**Figure 7:**
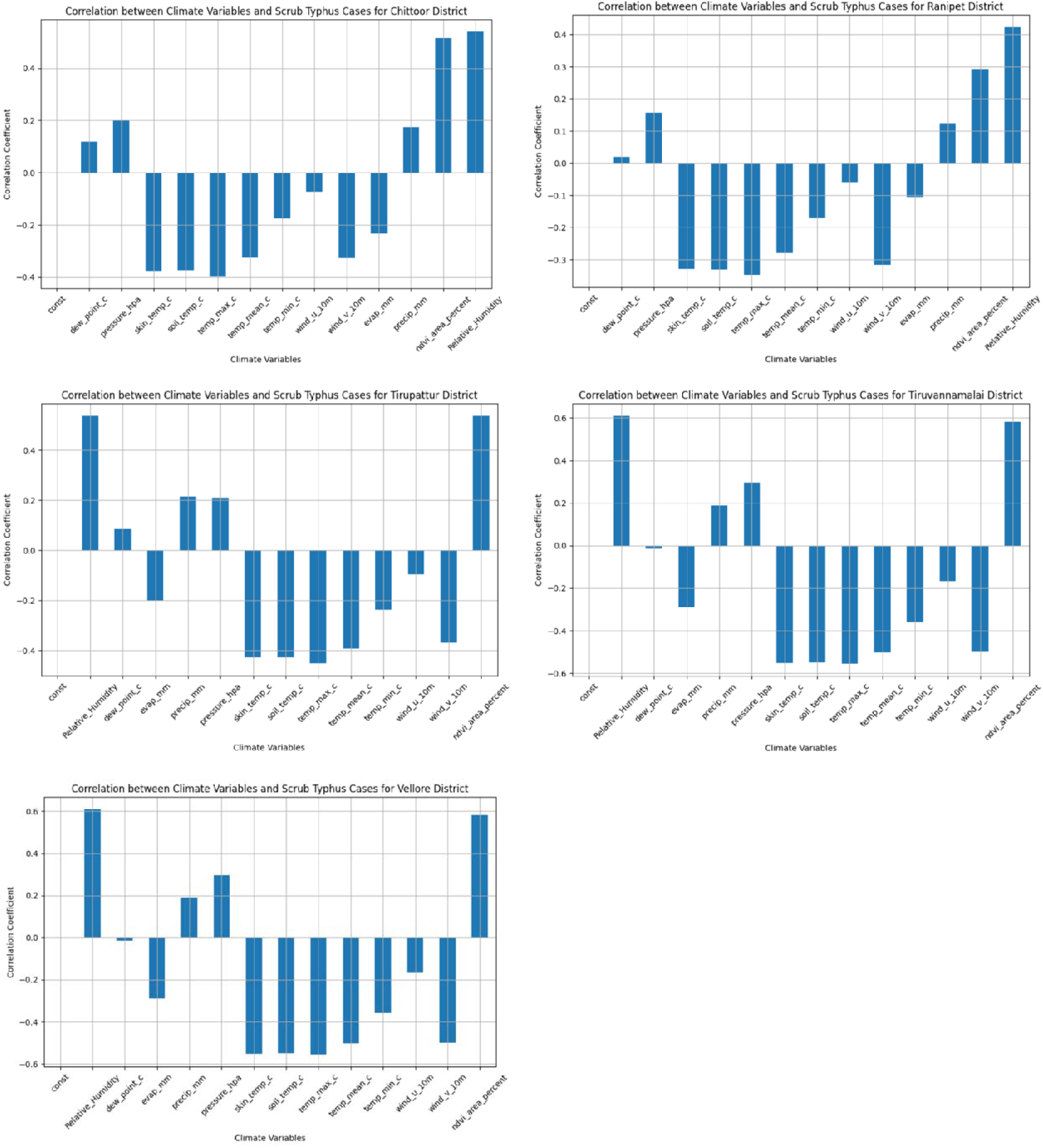
Significance of correlation from OLS regression analysis of scrub typhus risk factors across five study districts (May 2005–May 2024).

